# NLRP3 Activation and Impaired TGF-β Anti-Inflammatory Pathways Predict Vascular Risk in PWH on ART

**DOI:** 10.64898/2026.08.05.26359809

**Authors:** Alton S. Barbehenn, Caroline H. Sheikhzadeh, Sonia Savur, Erik Lundgren, Sannidhi Sarvadhavabhatla, Vivian Pae, Maria Sophia Donaire, Alejandro Schuler, Xiuping Chu, Colin T. Maguire, Senay Topal, Anuradha Ganesan, Joseph M. Yabes, Derek T. Larson, Tahaniyat Lalani, Evan C. Ewers, Rhonda E. Colombo, Jeffrey A. Tomalka, Priscilla Y. Hsue, Rafick P. Sekaly, Brian K. Agan, Sulggi A. Lee

## Abstract

**Importance:** The immune mechanisms driving vascular disease remain incompletely understood. People with HIV (PWH), even during effective antiretroviral therapy (ART), exhibit persistent immune activation and inflammation, which may contribute to higher rates of vascular disease and mortality compared with people without HIV (PWoH). Leveraging a cohort of U.S. military personnel followed from HIV diagnosis through long-term ART suppression, we sought to identify immunologic pathways underlying increased vascular risk.

**Objective:** To identify plasma biomarkers reflecting distinct immune mechanisms that predict incident vascular outcomes in ART-suppressed PWH.

**Design:** Case-cohort study within the U.S. Military HIV Natural History Study.

**Setting:** Longitudinal, multicenter observational cohort.

**Participants:** A total of 1,002 ART-suppressed PWH (HIV RNA <50 copies/mL) were included, with N=135 vascular event (VE) cases and N=702 controls. Cases encompassed atherosclerotic cardiovascular disease (ASCVD) – coronary artery disease (CAD), myocardial infarction (MI), stroke (CVA), peripheral artery disease (PAD) – and venous thrombotic events (VTE) – deep vein thrombosis (DVT) and pulmonary embolism (PE).

**Exposures:** Thirty-three soluble plasma analytes quantified using a high-sensitivity multiplex assay from samples collected ≥1 year after ART suppression.

**Main Outcomes and Measures:** The primary outcome was incident ASCVD. Associations between cytokine concentrations (individual and clustered) and vascular risk were evaluated using unsupervised clustering, Cox proportional hazards models, and causal inference (to estimate 5-year ASCVD risk under hypothetical cytokine alterations). Mediation analyses assessed direct and indirect effects of key inter-related cytokines. Secondary outcome included any VE (ASCVD plus VTE). Covariates included traditional cardiovascular risk factors, HIV clinical variables, and demographics. False discovery rate (FDR) adjustment was applied using the Benjamini-Hochberg method.

**Results:** Cytokine clusters reflecting NLRP3 inflammasome activation and persistent inflammation (IL-18, IL-6) and individual markers (IL-18: HR=1.89, q=0.007; TGF-β2: HR=0.74, q=0.026) were associated with increased ASCVD risk. IL-18 remained nominally significant after adjusting for traditional risk factors (p<0.05) but did not meet FDR significance (q<0.05).

**Conclusions and Relevance:** NLRP3 inflammasome activation and reduced TGF-β2, indicating loss of anti-inflammatory and repair mechanisms, may contribute to atherogenesis in ART-suppressed PWH. These findings highlight potential interventional targets for mitigating inflammation-driven vascular risk and warrant validation in larger cohorts to inform novel therapeutic strategies.

**Key Points:** *Question:* Which inflammatory immune pathways are associated with incident atherosclerotic cardiovascular disease in ART-suppressed people with HIV?

*Findings:* In this longitudinal case-cohort study of 1,002 ART-suppressed people with HIV, elevated IL-18 and reduced TGF-β2 were associated with incident ASCVD. Associations were specific to arterial rather than venous events, and IL-18-related risk was partly mediated through IL-6.

*Meaning:* These findings suggest that imbalance between inflammasome-driven inflammation and impaired immune regulation contributes to residual cardiovascular risk in treated HIV.

## INTRODUCTION

Vascular disease remains a leading cause of global morbidity and mortality.^1,2^ Atherosclerotic cardiovascular disease (ASCVD) – characterized by atherosclerotic plaque formation within arterial walls – is a major contributor to myocardial infarction (MI), coronary artery disease (CAD), ischemic cerebrovascular accidents (CVA), and peripheral artery disease (PAD). Despite advances in prevention and treatment, the immunologic mechanisms driving ASCVD remain incompletely defined.^3,4^

Systemic inflammation and immune activation are increasingly recognized as central drivers of vascular disease, contributing to both ASCVD and venous thromboembolism (VTE), including deep vein thrombosis (DVT) and pulmonary embolism (PE).^5–11^ Although ASCVD and VTE share inflammatory mechanisms, they arise through distinct pathophysiologic processes and may therefore have different immunologic determinants.^7–11^ These processes are amplified in people with HIV (PWH), even among those with durable viral suppression on antiretroviral therapy (ART).^12–14^ HIV infection is associated with increased risk of both ASCVD^12,15,16^ and VTE.^17^ Persistent immune activation, chronic inflammation, and endothelial dysfunction contribute to a prothrombotic state that persists despite ART.^18,19^ Because these immune perturbations can be measured early and longitudinally in select, well-characterized HIV cohorts, such studies provide a unique opportunity to identify upstream immunologic drivers of vascular disease with potential relevance to the general population.

Here, we analyzed plasma levels of 33 cytokines and chemokines in 1,002 ART-suppressed PWH from the U.S. Military HIV Natural History Study. We examined associations with incident ASCVD and overall vascular disease (including VTE) using clustering, survival modeling, and causal inference approaches. We specifically sought to identify early immune pathways that precede clinical disease. Our findings implicate NLRP3 inflammasome activation and impaired anti-inflammatory signaling via TGF-β as key and previously uncharacterized contributors to vascular risk, with potential relevance beyond HIV.

## METHODS

### Study Design and Sample Selection

A total of 1,002 PWH with sustained viral suppression (HIV-1 RNA <50 copies/mL for >1 year on ART) were selected from the U.S. Military HIV Natural History Study (NHS), a large, prospective, racially and ethnically diverse cohort with longitudinal follow-up, characterized by routine HIV screening and consistent diagnosis and care within the military healthcare system.^20^ Cardiovascular outcomes and relevant comorbidities in the NHS are systematically captured and adjudicated through structured participant interviews, detailed medical record review, and adjudication. We employed a case-cohort design including all eligible vascular event (VE) cases (N=200) and randomly selected controls (N=802) (**eFigure 1** in **Supplement 1**). VE was defined as incident ASCVD (CAD, MI, CVA, or PAD) and/or incident VTE (DVT or PE). Given the strong associations between age and vascular risk, patients at least 45 years old at HIV diagnosis were always included in the cohort. The resulting distributions of age, sex, and ethnicity were comparable between cases and controls (**Table 1**; **eTable 1**; **eFigure 2** in **Supplement 1**), and no additional sampling adjustments were performed. Eligible plasma samples (obtained between 1997 and 2015) were collected >1 year after ART suppression and at least one year prior to censorship or incident VE. To minimize reverse causation, the earliest eligible sample per participant was selected. After applying exclusion criteria, the final analytic sample included 135 VE cases (including 111 ASCVD cases) and 702 controls. This study was conducted and reported in accordance with the Strengthening the Reporting of Observational Studies in Epidemiology (STROBE) reporting guideline.

**Table 1.**
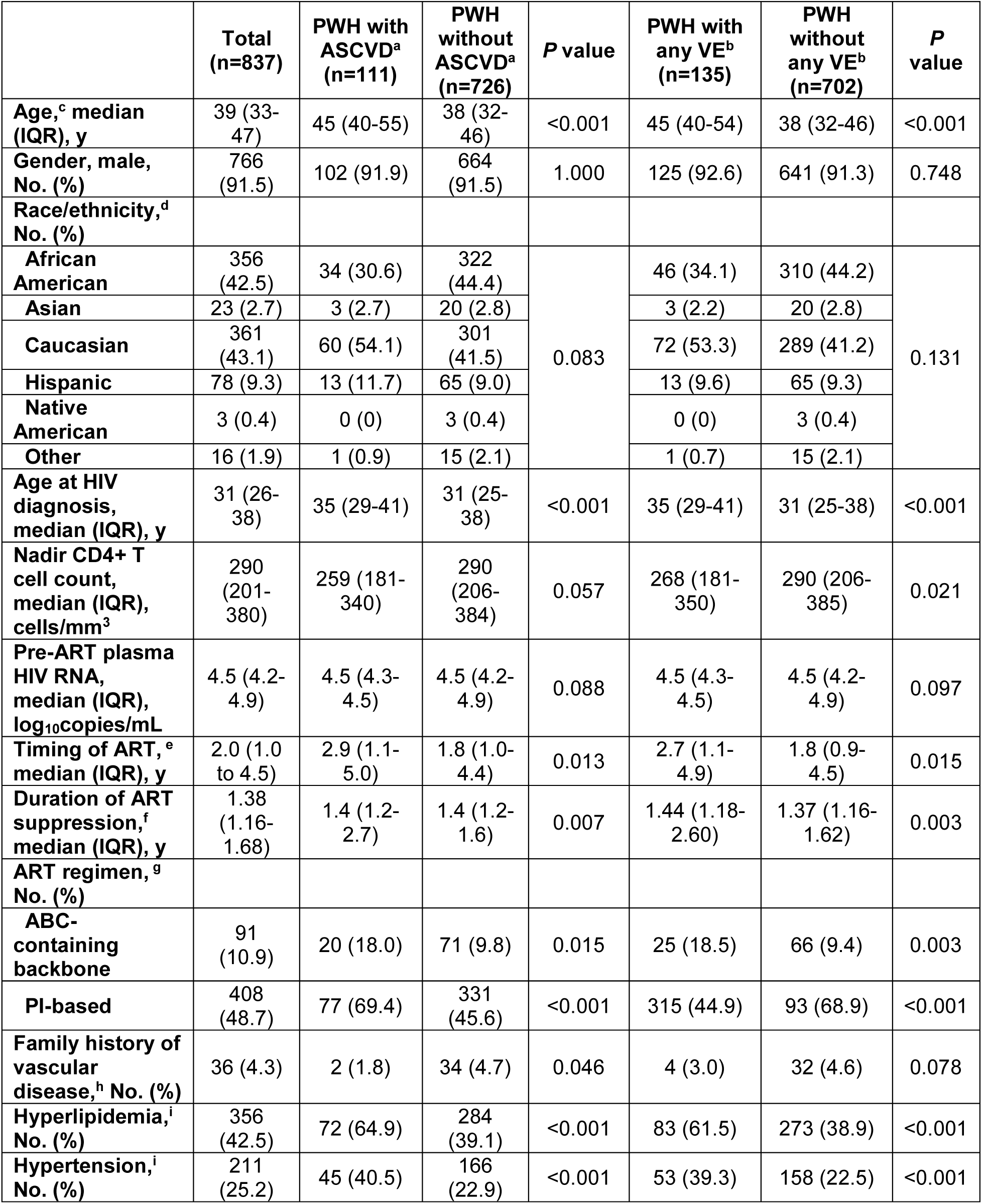
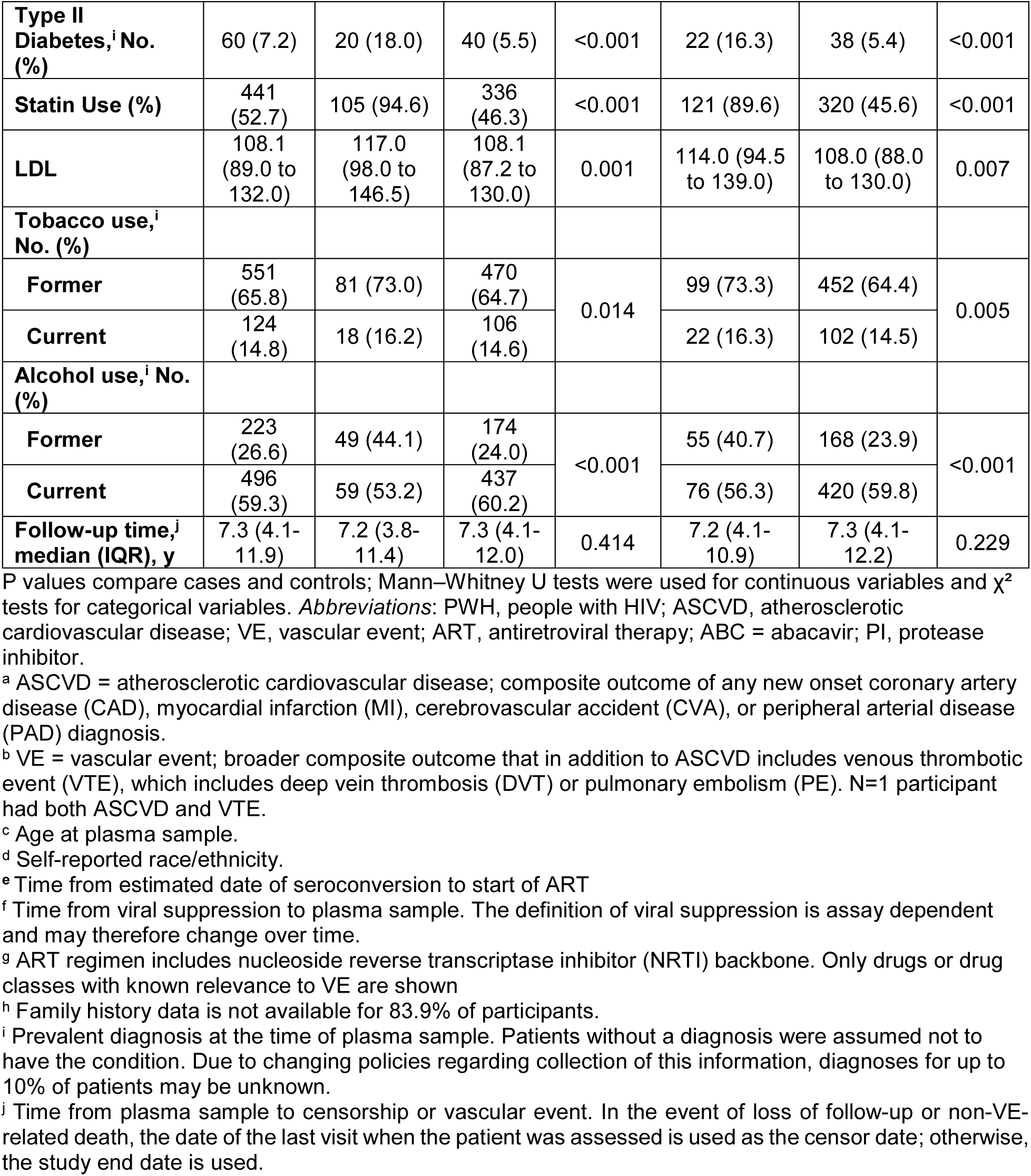
Baseline demographic and clinical characteristics of study participants.

|  | Total<br>(n=837) | PWH with<br>ASCVD<sup>a</sup><br>(n=111) | PWH<br>without<br>ASCVD<sup>a</sup><br>(n=726) | *P* value | PWH with<br>any VE<sup>b</sup><br>(n=135) | PWH<br>without<br>any VE<sup>b</sup><br>(n=702) | *P* value |
| --- | --- | --- | --- | --- | --- | --- | --- |
| Age,<sup>c</sup> median<br>(IQR), y | 39 (33-47) | 45 (40-55) | 38 (32-46) | <0.001 | 45 (40-54) | 38 (32-46) | <0.001 |
| Gender, male,<br>No. (%) | 766<br>(91.5) | 102 (91.9) | 664<br>(91.5) | 1.000 | 125 (92.6) | 641 (91.3) | 0.748 |
| Race/ethnicity,<sup>d</sup><br>No. (%) |  |  |  |  |  |  |  |
| African<br>American | 356<br>(42.5) | 34 (30.6) | 322<br>(44.4) | 0.083 | 46 (34.1) | 310 (44.2) | 0.131 |
| Asian | 23 (2.7) | 3 (2.7) | 20 (2.8) |  | 3 (2.2) | 20 (2.8) |  |
| Caucasian | 361<br>(43.1) | 60 (54.1) | 301<br>(41.5) |  | 72 (53.3) | 289 (41.2) |  |
| Hispanic | 78 (9.3) | 13 (11.7) | 65 (9.0) |  | 13 (9.6) | 65 (9.3) |  |
| Native<br>American | 3 (0.4) | 0 (0) | 3 (0.4) |  | 0 (0) | 3 (0.4) |  |
| Other | 16 (1.9) | 1 (0.9) | 15 (2.1) |  | 1 (0.7) | 15 (2.1) |  |
| Age at HIV<br>diagnosis,<br>median (IQR), y | 31 (26-38) | 35 (29-41) | 31 (25-38) | <0.001 | 35 (29-41) | 31 (25-38) | <0.001 |
| Nadir CD4+ T<br>cell count,<br>median (IQR),<br>cells/mm<sup>3</sup> | 290<br>(201-<br>380) | 259 (181-<br>340) | 290<br>(206-<br>384) | 0.057 | 268 (181-<br>350) | 290 (206-<br>385) | 0.021 |
| Pre-ART plasma<br>HIV RNA,<br>median (IQR),<br>log<sub>10</sub>copies/mL | 4.5 (4.2-4.9) | 4.5 (4.3-4.5) | 4.5 (4.2-4.9) | 0.088 | 4.5 (4.3-4.5) | 4.5 (4.2-4.9) | 0.097 |
| Timing of ART,<sup>e</sup><br>median (IQR), y | 2.0 (1.0<br>to 4.5) | 2.9 (1.1-5.0) | 1.8 (1.0-4.4) | 0.013 | 2.7 (1.1-4.9) | 1.8 (0.9-4.5) | 0.015 |
| Duration of ART<br>suppression,<sup>f</sup><br>median (IQR), y | 1.38<br>(1.16-1.68) | 1.4 (1.2-2.7) | 1.4 (1.2-1.6) | 0.007 | 1.44 (1.18-2.60) | 1.37 (1.16-1.62) | 0.003 |
| ART regimen,<sup>g</sup><br>No. (%) |  |  |  |  |  |  |  |
| ABC-containing<br>backbone | 91<br>(10.9) | 20 (18.0) | 71 (9.8) | 0.015 | 25 (18.5) | 66 (9.4) | 0.003 |
| PI-based | 408<br>(48.7) | 77 (69.4) | 331<br>(45.6) | <0.001 | 315 (44.9) | 93 (68.9) | <0.001 |
| Family history of<br>vascular<br>disease,<sup>h</sup> No. (%) | 36 (4.3) | 2 (1.8) | 34 (4.7) | 0.046 | 4 (3.0) | 32 (4.6) | 0.078 |
| Hyperlipidemia,<sup>i</sup><br>No. (%) | 356<br>(42.5) | 72 (64.9) | 284<br>(39.1) | <0.001 | 83 (61.5) | 273 (38.9) | <0.001 |
| Hypertension,<sup>i</sup><br>No. (%) | 211<br>(25.2) | 45 (40.5) | 166<br>(22.9) | <0.001 | 53 (39.3) | 158 (22.5) | <0.001 |
| **Type II Diabetes,<sup>i</sup> No. (%)** | 60 (7.2) | 20 (18.0) | 40 (5.5) | <0.001 | 22 (16.3) | 38 (5.4) | <0.001 |
| **Statin Use (%)** | 441 (52.7) | 105 (94.6) | 336 (46.3) | <0.001 | 121 (89.6) | 320 (45.6) | <0.001 |
| **LDL** | 108.1 (89.0 to 132.0) | 117.0 (98.0 to 146.5) | 108.1 (87.2 to 130.0) | 0.001 | 114.0 (94.5 to 139.0) | 108.0 (88.0 to 130.0) | 0.007 |
| **Tobacco use,<sup>i</sup> No. (%)** |  |  |  |  |  |  |  |
| **Former** | 551 (65.8) | 81 (73.0) | 470 (64.7) | 0.014 | 99 (73.3) | 452 (64.4) | 0.005 |
| **Current** | 124 (14.8) | 18 (16.2) | 106 (14.6) |  | 22 (16.3) | 102 (14.5) |  |
| **Alcohol use,<sup>i</sup> No. (%)** |  |  |  |  |  |  |  |
| **Former** | 223 (26.6) | 49 (44.1) | 174 (24.0) | <0.001 | 55 (40.7) | 168 (23.9) | <0.001 |
| **Current** | 496 (59.3) | 59 (53.2) | 437 (60.2) |  | 76 (56.3) | 420 (59.8) |  |
| **Follow-up time,<sup>j</sup> median (IQR), y** | 7.3 (4.1-11.9) | 7.2 (3.8-11.4) | 7.3 (4.1-12.0) | 0.414 | 7.2 (4.1-10.9) | 7.3 (4.1-12.2) | 0.229 |

P values compare cases and controls; Mann–Whitney U tests were used for continuous variables and $\chi^2$ tests for categorical variables. *Abbreviations:* PWH, people with HIV; ASCVD, atherosclerotic cardiovascular disease; VE, vascular event; ART, antiretroviral therapy; ABC = abacavir; PI, protease inhibitor.
<sup>a</sup> ASCVD = atherosclerotic cardiovascular disease; composite outcome of any new onset coronary artery disease (CAD), myocardial infarction (MI), cerebrovascular accident (CVA), or peripheral arterial disease (PAD) diagnosis.
<sup>b</sup> VE = vascular event; broader composite outcome that in addition to ASCVD includes venous thrombotic event (VTE), which includes deep vein thrombosis (DVT) or pulmonary embolism (PE). N=1 participant had both ASCVD and VTE.
<sup>c</sup> Age at plasma sample.
<sup>d</sup> Self-reported race/ethnicity.
<sup>e</sup> Time from estimated date of seroconversion to start of ART
<sup>f</sup> Time from viral suppression to plasma sample. The definition of viral suppression is assay dependent and may therefore change over time.
<sup>g</sup> ART regimen includes nucleoside reverse transcriptase inhibitor (NRTI) backbone. Only drugs or drug classes with known relevance to VE are shown
<sup>h</sup> Family history data is not available for 83.9% of participants.
<sup>i</sup> Prevalent diagnosis at the time of plasma sample. Patients without a diagnosis were assumed not to have the condition. Due to changing policies regarding collection of this information, diagnoses for up to 10% of patients may be unknown.
<sup>j</sup> Time from plasma sample to censorship or vascular event. In the event of loss of follow-up or non-VE-related death, the date of the last visit when the patient was assessed is used as the censor date; otherwise, the study end date is used.

### Plasma cytokine quantification

Cryopreserved plasma samples were analyzed using a custom U-Plex multiplex chemiluminescent assay (MesoScale Discovery, Rockland, MD), using a panel previously developed and validated in related studies.^21–24^ Concentrations of 33 cytokines and chemokines were measured (**eMethods**; **eTable 2** in **Supplement 1**). Cytokines with >15% missingness were excluded from downstream analyses. Batch effects were addressed using ComBat on log-scaled concentrations.^25^

### Statistical Analysis

Cytokine concentrations were compared between case and control groups using Mann-Whitney tests. Cytokine cluster medoids were created by first log-scale normalizing each cytokine across participants (mean zero, variance one), and the cluster median was then computed within participant.

Cox proportional hazard (CPH) models were used to estimate the relationship between cytokines and the vascular event hazard rate, with time measured from one year after plasma sample collection. Stratified Barlow weights were used to fit CPH models to account for the stratified case-cohort study design (different sampling probabilities differed by age of documented HIV seroconversion).^26,27^ Censoring was assumed to be non-informative. A cubic b-spline with four degrees of freedom and regular knots was used in nonlinear models of cytokine concentration. All cytokine modeling was performed on a log_2_-scale. The Benjamini-Hochberg procedure was used for false discovery rate correction.

Covariates for multivariate modeling included ASCVD risk factors (age at plasma sample, sex, race/ethnicity, existing diagnosis of hypertension, hyperlipidemia, and type II diabetes, family history of cardiovascular disease, and tobacco use) and HIV clinical variables (nadir CD4+ T cell count, log_10_ pre-ART viral load, timing of ART initiation, duration of ART suppression, and ART regimen).

Counterfactual 5-year ASCVD risk (among patients with no VE within one year after the plasma sample) was estimated under a multiplicative modified treatment policy (MTP) corresponding to a 2-fold increase in individual plasma cytokine concentrations.^28–31^ The counterfactual ASCVD risk was further decomposed using a machine-learning mediation estimator,^32^ framed as a point-treatment classification problem with a single intervention time. Counterfactual models were adjusted for every covariate considered for inclusion in the multivariate CPH modeling. Both logistic regression and random forest learners were used to produce the point-estimates of counterfactual risk.

Statistical analyses were conducted in R (v4.5.3) using the following packages: survival (v3.8-6), survey (v4.5), lmtp (v1.4.0), and crumble (v0.1.2).

## RESULTS

### Characteristics of study participants

Cases and controls were broadly similar with respect to sex, race/ethnicity, pre-ART viral load, and nadir CD4+ T-cell count (**Table 1**). Cases were slightly older, had longer time to ART initiation and duration of viral suppression, and had a higher prevalence of type II diabetes, hyperlipidemia, and hypertension at the time of plasma collection. These variables were included as covariates in multivariable analyses.

### Plasma cytokine values varied considerably by assay detection levels and demonstrated batch effects

Detectability and dynamic range varied across the 33 analytes (**eTable 2** in **Supplement 1**), with some cytokines demonstrating low detection consistent with known assay limitations (e.g., IL-1β), whereas others (e.g., IL-6 and IL-18) were reliably quantified. Samples were processed in two batches; however, variability across assay plates was observed (**eFigure 3** and **eFigure 4A** in **Supplement 1**). To ensure data reliability, samples collected within one week from the same individual were treated as biological replicates; discordant replicates (p<0.05) were excluded, and concordant values were averaged. After quality control and exclusion of cytokines with >15% missingness, 20 analytes remained for analysis (fractalkine, I-TAC, IFN-γ, IL-10, IL-15, IL-18, IL-1β, IL-27, IL-29, IL-6, IL-7, IL-8, IL-9, IP-10, MCP-3, MIP-1α, MIP-3α, TGF-β1, TGF-β2, and TNF-α), with improved distributional consistency across plates after batch correction (**eFigure 4B** in **Supplement 1).**

### Cytokine correlation structure during ART suppression

Unsupervised hierarchical clustering of plasma cytokines identified coordinated patterns of immune activation among analytes measured after ≥1 year of ART suppression (**Figure 1**). Six clusters were observed, broadly reflecting distinct but overlapping immune pathways.

**Figure 1.**
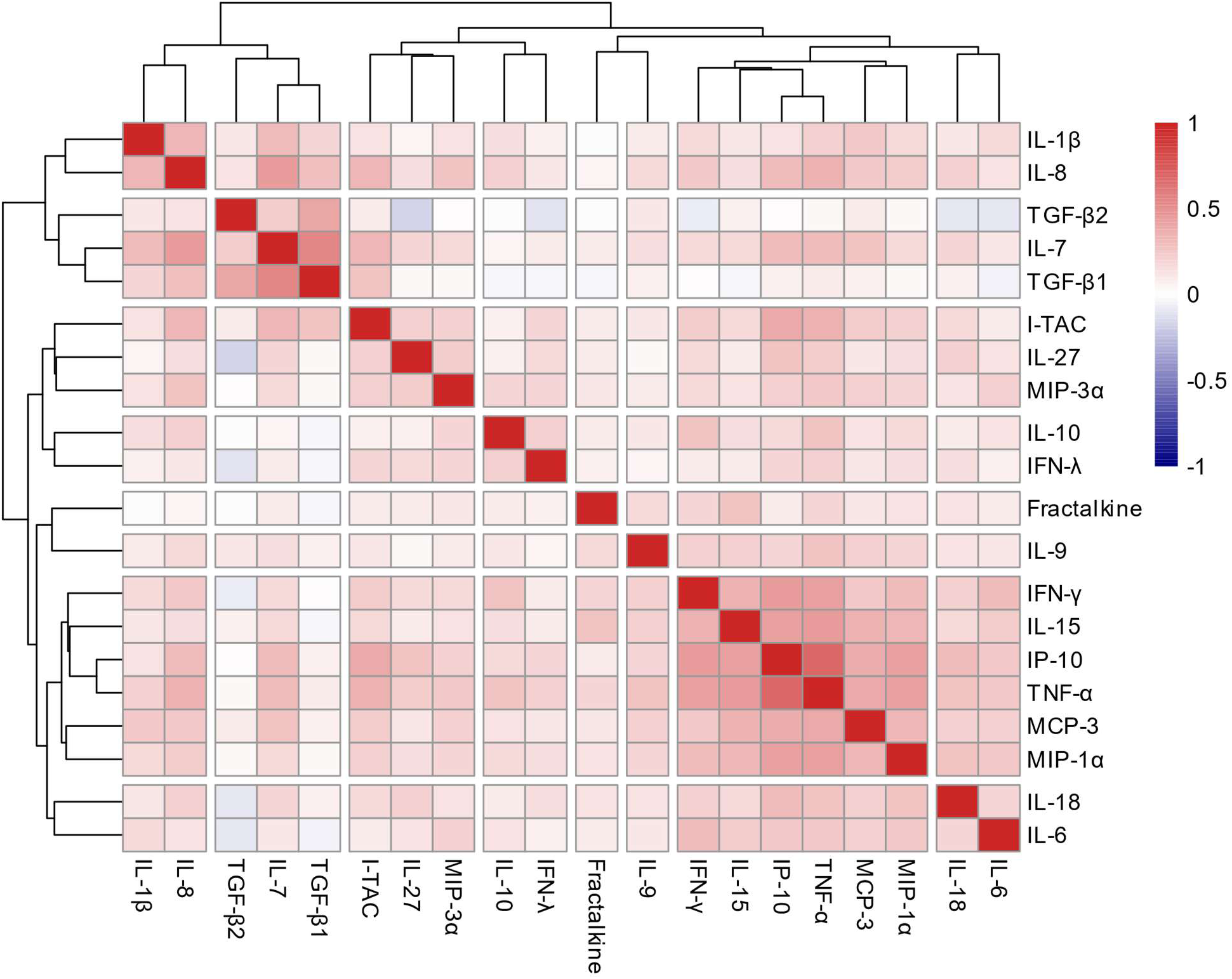
Plasma cytokine concentrations after approximately one year of ART-mediated viral suppression. The heatmap displays Spearman correlation coefficients (ρ) between pairs of cytokines. Cytokines were ordered using hierarchical clustering (Ward’s method) based on the distance metric 1 − |*ρ*|, such that cytokines with strong positive or negative correlations are grouped together.

Cluster 1 included IFN-γ, IL-15, IP-10, TNF-α, MCP-3, and MIP-1α, consistent with Th1-polarized and macrophage-associated inflammatory responses. Cluster 2 comprised IL-18 and IL-6, reflecting systemic inflammation and pathways linked to inflammasome activity. Cluster 3 (IL-1β, IL-8) was consistent with innate immune activation and neutrophil recruitment. Cluster 4 (IL-7, TGF-β1, TGF-β2) aligned with immune homeostasis, lymphocyte maintenance, and tissue repair processes. Cluster 5 (I-TAC, IL-27, MIP-3α) reflected immune activation with chemotactic and regulatory signaling. Cluster 6 (IL-10, IL-29) was consistent with immunoregulatory and antiviral signaling pathways. Fractalkine and IL-9 did not cluster strongly with other analytes, suggesting more independent or context-specific roles in immune signaling.

### Plasma IL-18 and TGF-β2 levels were significantly different in ASCVD cases than controls

Among quality-controlled cytokines, several differed between ASCVD cases and controls (**Figure 2**). IL-18 levels were significantly higher in ASCVD cases (Mann-Whitney test, p<0.001), with IP-10 also elevated (p<0.01). Additional cytokines showed nominal differences, including higher IL-6, IFN-γ, IL-27, IL-8, MIP-1α, TNF-α, and IL-15, and lower TGF-β2 (all p<0.05). However, after correction for multiple comparisons, only IL-18 and IP-10 remained significant (q=0.015 and 0.049, respectively). Median IL-18 and IP-10 concentrations were approximately 12% and 10% higher in cases than controls.

**Figure 2.**
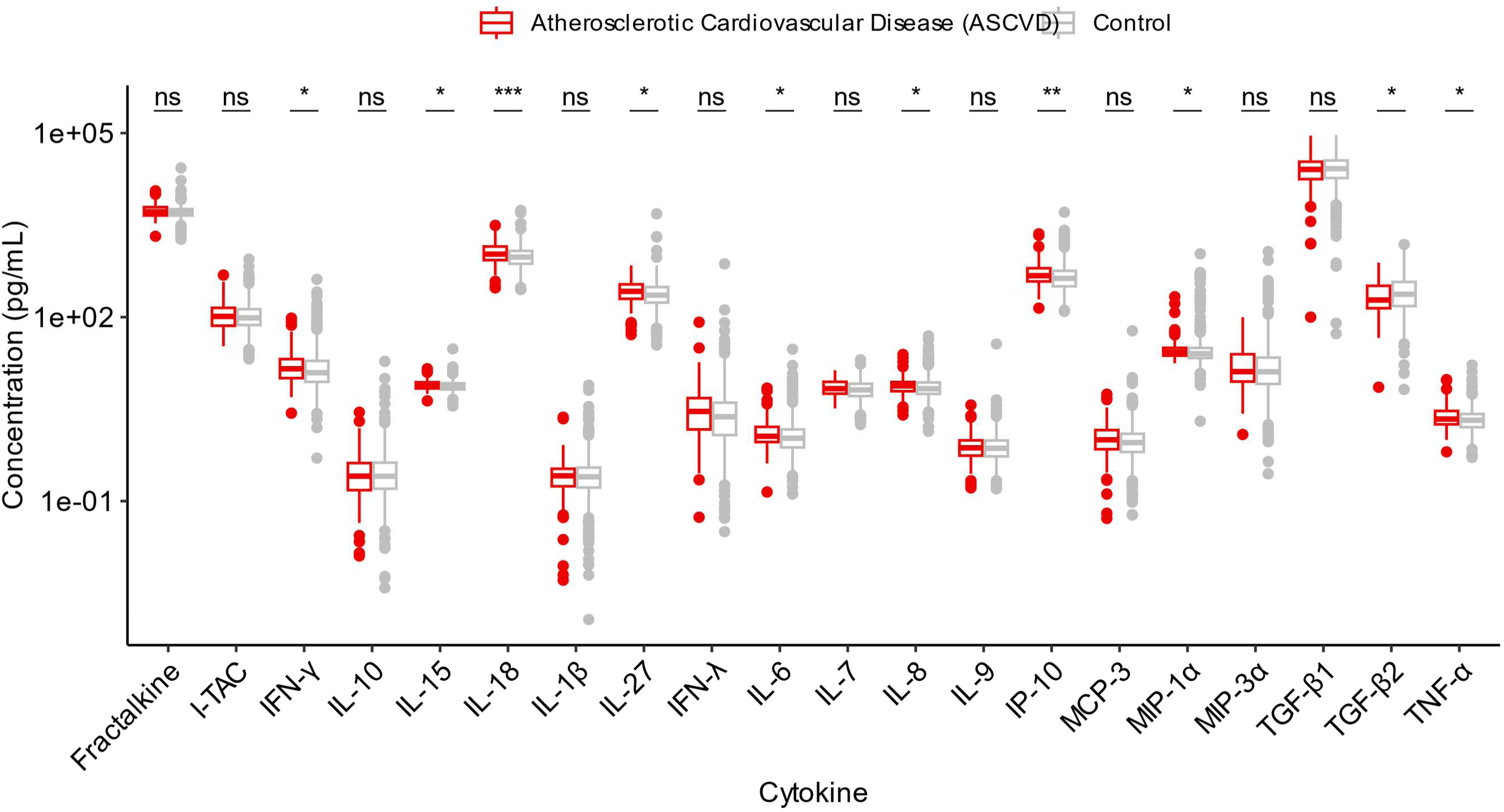
Comparison of plasma cytokine concentrations between ASCVD cases and controls. This figure compares plasma cytokine concentrations between ASCVD cases and controls using Mann-Whitney U tests. Significance levels are indicated as follows: *** for p <0.001, ** for p <0.01, * for p <0.05, and ‘ns’ for p ≥0.05. All statistically significant cytokines (p <0.05), except TGF-β2, were elevated in ASCVD cases compared to controls. After false discovery rate (FDR) correction using the Benjamini-Hochberg procedure, only IL-18 (q=0.014) and IP-10 (q=0.049) remained statistically significant (q <0.05).

Across ASCVD subtypes, IL-18 was consistently elevated in cases (CAD, MI, CVA, and PAD), whereas IP-10 elevations were primarily observed in CAD and MI (**eFigure 5** in **Supplement 1**). In contrast, neither IL-18 nor IP-10 differed significantly in VTE. When ASCVD and VTE were combined into a composite vascular endpoint, associations were attenuated (**eFigure 6** in **Supplement 1**).

### Plasma cytokines, particularly IL-18 and TGF-β2, were associated with incident ASCVD

Associations between baseline plasma cytokines and incident ASCVD were evaluated using Cox proportional hazards models, with cytokine concentrations log_2_-transformed to estimate hazard ratios per two-fold increase. ASCVD was analyzed separately from VTE given distinct pathophysiologic mechanisms.^33,34^

In unadjusted analyses, five cytokines were associated with ASCVD (all p<0.05), but only IL-18 and TGF-β2 remained significant after false discovery rate correction (both q<0.05) (**Figure 3**; **eTable 3A** in **Supplement 1**). IL-18, a key known pro-inflammatory cytokine that drives atherosclerotic plaque instability,^35^ showed the strongest positive association (HR=1.89, 95% CI=1.33-2.68, p=3.5e-4), while TGF-β2, a known immunoregulatory cytokine,^36,37^ was inversely associated with risk (HR=0.74, 95% CI=0.61-0.90). Other cytokines, including IL-15 (HR=1.92, CI=1.08-3.43, p=0.026), TNF-α (HR=1.43, CI=1.06-1.93, p=0.020), and IP-10 (HR=1.38, CI=1.05-1.81, p=0.020) showed nominal but non-FDR-significant associations.

**Figure 3.**
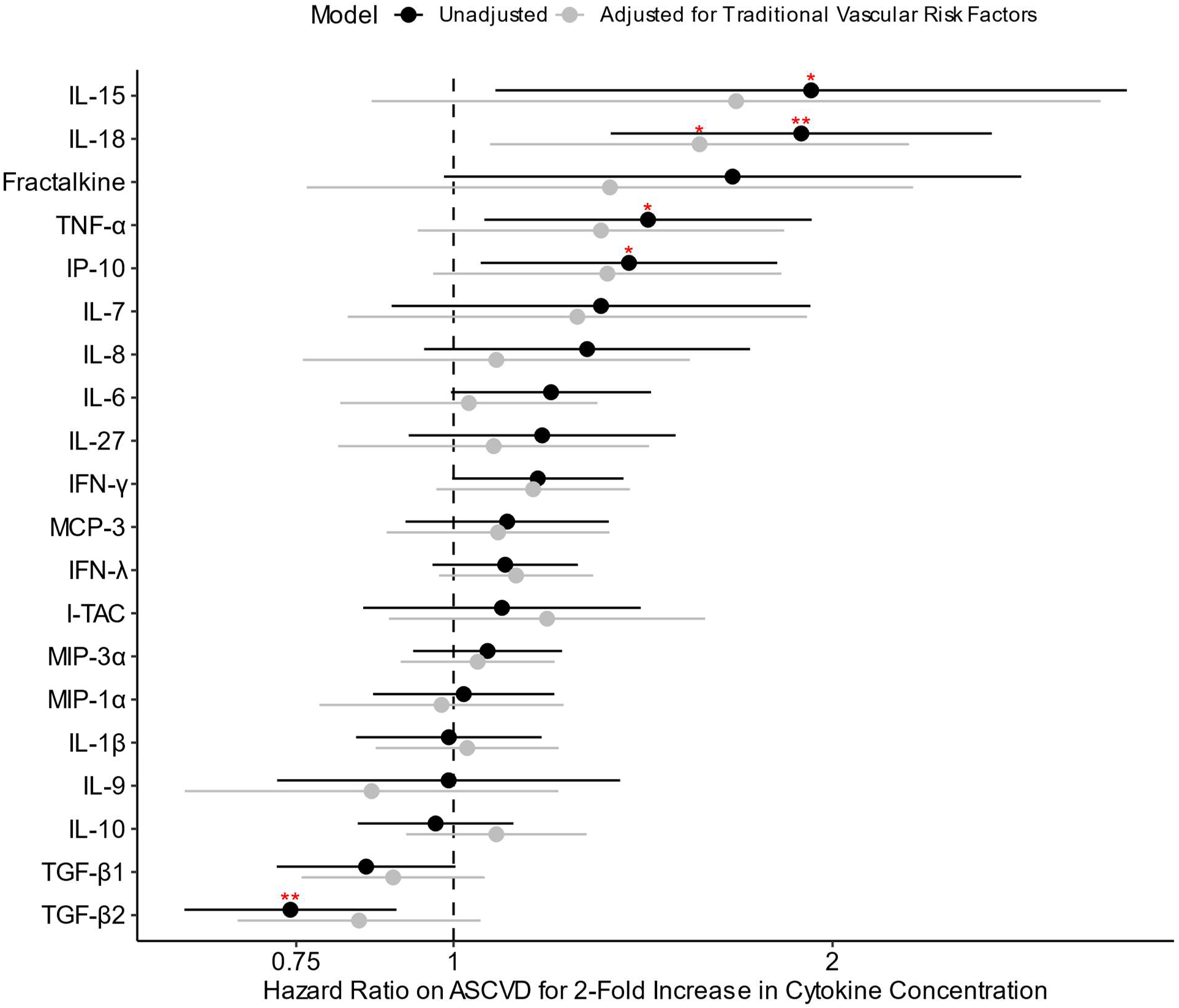
Unadjusted and adjusted hazard ratios of ASCVD for a 2-fold increase in plasma cytokine concentration. A linear unadjusted (black) and adjusted (grey) Cox proportional hazard models were fit to each cytokine individually. Adjusted models control for traditional vascular risk factors - age (at plasma sample), sex, race/ethnicity, family history of vascular disease, tobacco use, and comorbidities (diagnosis of hyperlipidemia, hypertension, or type II diabetes). Cytokines found to be significant at a nominal p-value level (p < 0.05) or at a false discovery rate corrected level (q < 0.05) are denoted * and **, respectively. False discovery rate correction was applied to each model separately. Bars represent 95% confidence intervals for each hazard ratio estimate using robust standard error.

After adjustment for demographic and cardiovascular risk factors, IL-18 remained nominally associated with ASCVD risk but did not meet FDR significance (aHR=1.57, 95% CI=1.07-2.30, p=0.02), whereas TGF-β2 remained directionally consistent but was not statistically significant (**Figure 3**; **eTable 3A** in **Supplement 1**). Further adjustment for HIV-related clinical variables did not materially alter these estimates (**eFigure 7** in **Supplement 1**).

Sensitivity analyses using cubic B-splines supported an approximately linear dose-response relationship for IL-18, with weaker evidence for nonlinearity for TGF-β2 (q=0.033 and 0.069, respectively) (**eTable 3B** in **Supplement 1**). When VTE was included in a composite vascular endpoint, effect estimates were attenuated; however, IL-18 remained FDR-significant (HR=1.67, q=0.035), and TGF-β2 remained directionally consistent and nominally significant (HR=0.80, p=0.013) (**eFigure 8** and **eTable 4** in **Supplement 1**). With additional adjustment for demographic and cardiovascular risk factors, neither association remained statistically significant. At the cluster level (**eFigure 9** in **Supplement 1**), cytokines in Cluster 3 (IFN-γ, IL-15, IP-10, MCP-3, MIP-1α, TNF-α) and Cluster 5 (IL-18 and IL-6) – which included several cytokines that were individually predictive (**Figure 3**) – were significantly associated with ASCVD risk at an FDR-adjusted q<0.05 (**eTable 5** in **Supplement 1**). However, after further adjustment for traditional risk factors, these associations were no longer statistically significant.

### Causal modeling analyses indicated a potential role for IL-18 in ASCVD risk

Following identification of IL-18 and TGF-β2 as the strongest correlates of ASCVD in Cox proportional hazards models, we applied modified treatment policy estimation to evaluate their potential causal effects on 5-year ASCVD risk. Models adjusted for traditional cardiovascular risk factors and HIV-related clinical characteristics, without inclusion of other cytokines as covariates. Of the 2 cytokines evaluated in causal inference analyses, neither was significantly associated with ASCVD risk. However, the estimated effect of IL-18 corresponded to a relative risk (RR) of 2.12 (95% CI 0.67–6.77, p=0.203), whereas TGF-β2 was associated with a more modest estimated increase in risk (RR=1.22, 95% CI 0.89–1.68, p=0.214). (**eFigure 10**; **eTable 6** in **Supplement 1**). Given shared NLRP3 inflammasome-related signaling between IL-18 and IL-1β, and the use of IL-18 as a circulating proxy of inflammasome activation, we performed a sensitivity analysis additionally adjusting for IL-1β. The estimated effect of IL-18 remained similar after adjustment (RR=2.31, 95% CI=0.85-6.33, p=0.103), suggesting robustness of the observed association and supporting a potential role for IL-18–related inflammasome signaling independent of IL-1β.

### IL-18-associated ASCVD risk is partially mediated by IL-6

IL-18 and IL-1β are both released upon NLRP3 inflammasome activation and independently induce IL-6 production,^38^ a cytokine strongly associated with incident ASCVD in the general population.^39,40^ To evaluate whether IL-18 was associated with ASCVD beyond these related inflammatory mediators, we constructed Cox proportional hazards models with mutual adjustment for IL-18, IL-1β, IL-6, and standard cardiovascular risk factors. IL-18 remained significantly associated with ASCVD risk after adjustment for IL-1β and IL-6 (HR=1.54, 95% CI=1.03-2.31, p=0.036; **eFigure 11** in **Supplement 1**).

We then applied modified treatment policy estimation to assess the effect of a two-fold increase in IL-18 on ASCVD risk, partitioning effects into direct and IL-6-mediated pathways based on an externally validated causal structure (**Figure 4**; **eTable 7** in **Supplement 1**). After adjustment for conventional cardiovascular and HIV-related clinical variables, approximately 75% of the total IL-18 effect was mediated through IL-6. Both the direct (risk increase [RI]=0.015, 95% CI=0.003-0.027) and indirect (RI=0.043, 95% CI=0.036-0.049) effects were statistically significant (p<0.05), suggesting that IL-6 contributes substantially but does not fully account for IL-18-associated ASCVD risk. Incorporation of IL-1β as an additional confounder increased both the direct (RI=0.014 vs 0.026 with adjustment) and IL-6-mediated (RI=0.043 vs 0.073) effects of IL-18, consistent with negative confounding. These findings suggest that failure to account for IL-1β may underestimate the contribution of IL-18–driven inflammasome signaling to ASCVD risk.

**Figure 4.**
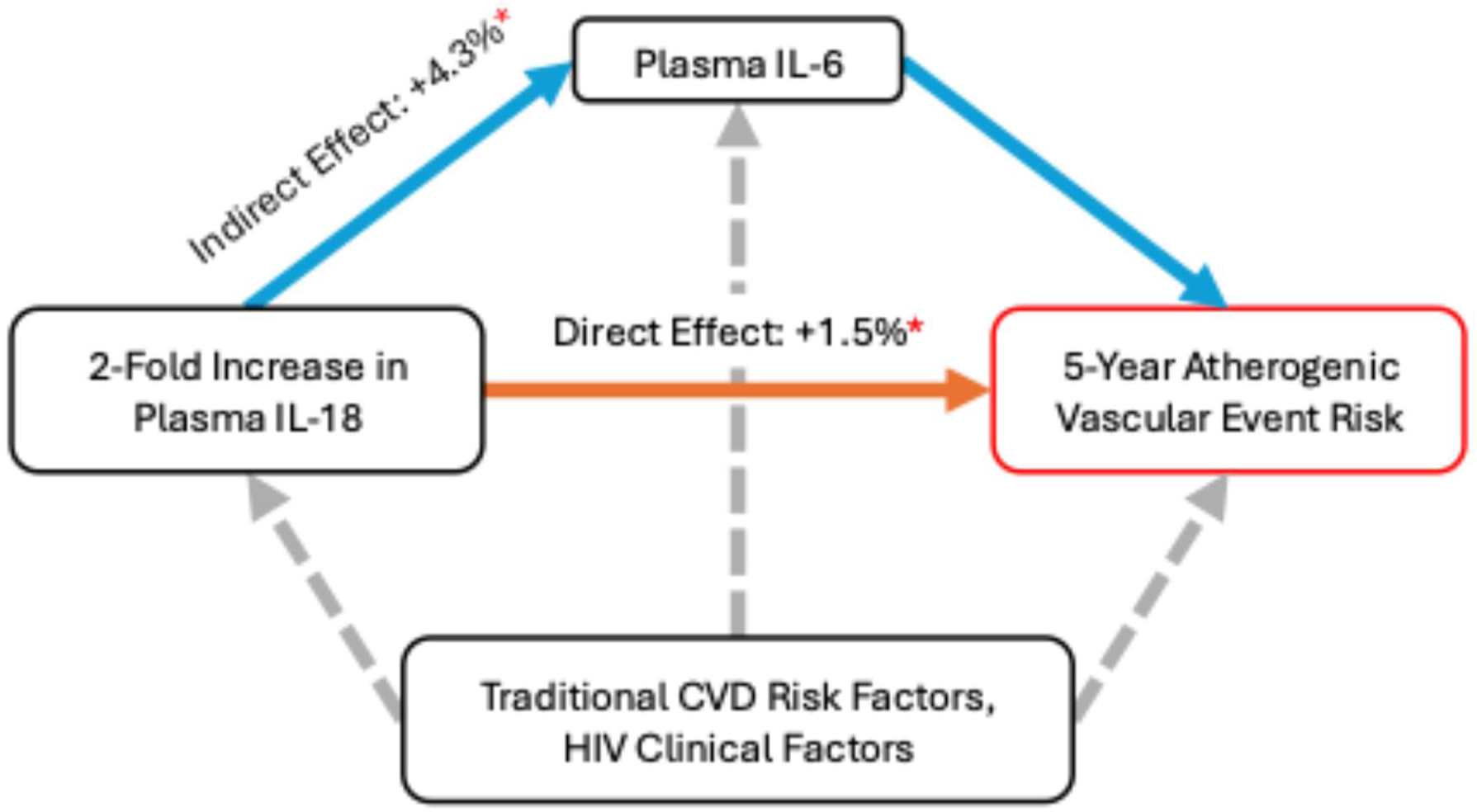
Higher levels of IL-18 increase 5-year ASCVD risk primarily mediated through IL-6 in causal inference models. A directed acyclic graph depicting the hypothesized causal pathway by which IL-18 might affect 5-year ASCVD risk is depicted. Both the direct effect (orange arrows) and indirect effect through IL-6 (blue arrows) are adjusted for traditional ASCVD risk factors (age, sex, race/ethnicity, family history of vascular disease, tobacco use, comorbidities (diagnosis of hyperlipidemia, hypertension, or type II diabetes), and HIV clinical factors (timing of ART initiation, duration of viral suppression, nadir CD4+ T count, pre-ART viral load, and ART regimen (given known effects of protease inhibitors on lipids and abacavir on NLRP3 inflammasome)) (dashed grey arrows). Absolute risk increases estimated for each pathway are labeled on the figure; a red asterisk indicates statistical significance (p < 0.05). *Abbreviations*: CVD, cardiovascular disease.

## DISCUSSION

In this longitudinal study of ART-suppressed PWH, we identified immunologic signatures associated with incident ASCVD despite durable viral suppression. Elevated IL-18 and reduced TGF-β2 implicated persistent inflammasome activation and impaired immunoregulatory signaling as key pathways underlying residual vascular risk. Cluster-based analyses reinforced these findings, suggesting coordinated inflammatory and regulatory immune networks contribute to ASCVD risk. These results extend prior work by demonstrating that immune perturbations measured during clinically stable ART suppression are associated with subsequent vascular events.

A central finding of this study is the robust association between IL-18 and incident ASCVD. IL-18 is a downstream effector of NLRP3 inflammasome activation and has been linked to plaque destabilization, endothelial dysfunction, and vascular inflammation. Our findings are consistent with prior evidence implicating inflammasome signaling in atherosclerosis, including the CANTOS trial, in which IL-1β inhibition reduced recurrent cardiovascular events independent of lipid lowering,^41^ and our prior study demonstrating that canakinumab reduced arterial inflammation in ART-treated PWH.^42^ Because IL-18 and IL-1β are co-regulated through NLRP3 activation,^38^ IL-18 may serve as a more stable circulating marker of upstream inflammasome activity than IL-1β, which is difficult to quantify reliably in plasma. Although the association was attenuated after adjustment for traditional cardiovascular risk factors, randomized clinical trials demonstrating a 31% reduction in the composite risk of cardiovascular death, myocardial infarction, ischemic stroke, or coronary revascularization with colchicine further support the NLRP3 inflammasome as a clinically relevant therapeutic target.^43^

A key observation in this study is the phenotype specificity of these associations. IL-18 was consistently elevated across ASCVD subtypes. When arterial and venous outcomes were combined (VE), associations were attenuated, suggesting that inflammasome-associated cytokine signaling may be more linked to arterial than venous thrombosis, consistent with distinct biological mechanisms governing these vascular beds.^33,34^

Causal mediation analyses suggested that approximately 75% of the IL-18-associated ASCVD risk was mediated through IL-6, consistent with prior genetic and clinical studies implicating IL-6 signaling in cardiovascular disease.^44–49^ Our findings extend those of Reilly et al.^49^ by placing IL-18 upstream of IL-6 while demonstrating that IL-6 accounts for only part of the associated cardiovascular risk. Persistence of a direct IL-18 effect independent of IL-6 suggests additional mechanisms, such as endothelial activation^50^ and pyroptosis.^38,51^ Together, these findings provide a rationale for therapeutic strategies targeting inflammasome signaling upstream of IL-6, an approach currently under investigation in clinical trials.^52,53^

Reduced TGF-β2 was independently associated with ASCVD, identifying impaired immunoregulatory signaling as a complementary mechanism contributing to vascular risk. TGF-β signaling regulates immune homeostasis, vascular integrity, and tissue repair, and diminished activity has been associated with plaque instability and vascular inflammation.^54,55^ These findings suggest that residual cardiovascular risk reflects not only persistent inflammation but also impaired resolution and repair.

These findings should be interpreted in the context of several limitations. First, although this was a relatively large and well-characterized HIV cohort, the number of ASCVD events limited statistical power, particularly after correction for multiple comparisons. This challenge is common across longitudinal HIV cohorts, which were established early in the epidemic and often include relatively young participants with insufficient follow-up to accrue large numbers of cardiovascular events.^56–59^ Consequently, composite vascular outcomes are frequently used to improve statistical power,^60–62^ although our findings highlight important biological differences between arterial and venous disease. Second, although we applied rigorous causal inference methods, residual confounding cannot be excluded, including the use of binary measures of hypertension, hyperlipidemia, and diabetes that do not account for disease severity, duration, or treatment. Third, cytokines were measured at a single time point after ART suppression and therefore may not capture longitudinal immune dynamics. Finally, the cohort consisted predominantly of men and included participants receiving older ART regimens, including abacavir and protease inhibitors, which may limit generalizability to contemporary HIV populations. Despite these limitations, strengths include the longitudinal design, precise ART and sampling timing, rigorous biomarker quantification with batch correction, and integration of clustering, survival, and causal mediation analyses.

In conclusion, among ART-suppressed people with HIV, elevated IL-18 and reduced TGF-β2 were associated with incident ASCVD. Collectively, these findings support a model in which chronic immune dysregulation – characterized by persistent inflammasome activation, downstream IL-6 signaling, and impaired TGF-β-mediated immunoregulation – may contribute to residual cardiovascular risk despite effective ART. These results identify upstream inflammasome signaling as a potential therapeutic target for cardiovascular risk reduction beyond traditional risk factor modification and warrant validation in larger, more diverse cohorts.

## Supporting information

Supplementary Online Content

## Data Availability

Data underlying the findings reported in this article are available from the corresponding author upon reasonable request, subject to institutional review and approval and completion of a data use agreement. Deidentified participant data, along with a data dictionary, will be made available to qualified investigators for purposes of replicating procedures and results.

## ACKNOWLEDGEMENTS

The authors wish to acknowledge the participation of all the study participants who contributed to this work as well as the clinical research staff of the U.S. Military HIV Natural History Study cohort who made this research possible. This work was supported in part by the National Institutes of Health: K23GM112526 (SAL), R01AI143464 (SAL), and UM1TR004409 (CTM). Partial support for this work (IDCRP-000-03) was provided by the Infectious Disease Clinical Research Program (IDCRP), a Department of Defense program executed through the Uniformed Services University of the Health Sciences, Department of Preventive Medicine and Biostatistics through a cooperative agreement with The Henry M. Jackson Foundation for the Advancement of Military Medicine, Inc. (HJF). This project has been funded in part by the National Institute of Allergy and Infectious Diseases, National Institutes of Health, under Inter-Agency Agreement Y1-AI-5072, and the Defense Health Program, U.S. DoW, under award HU0001190002. The views expressed are those of the authors and do not reflect the official views of the Uniformed Services University of the Health Sciences, the Henry M. Jackson Foundation for the Advancement of Military Medicine, Inc., the National Institutes of Health or the Department of Health and Human Services, the Department of Defense, Defense Health Agency, the Departments of the Army, Navy or Air Force, and U.S. Government. The funders had no role in study design, data collection and analysis, decision to publish, or preparation of the manuscript.

## AUTHOR CONTRIBUTIONS STATEMENT

All authors provided critical feedback in finalizing the report. SAL conceived, designed, and obtained funding to support the study with critical feedback from PYH, RPS, and BKA. BKA and XC coordinated the collection, management, and quality control processes for the clinical data, and BKA, ST, AG, JMY, DTL, TL, ECE, and REC provided the biospecimens. BKA, XC, SS, VP, MSD, CHS, SoS, CTM, and SAL managed the shipment and processing of samples. JAT and RPS performed the cytokine assays. ASB, SS, VP, MSD, CHS, SoS, BKA, XC, and SAL performed quality control analyses of the clinical data, and JAT, ASB, and SAL performed quality control analyses of the cytokine assay data. ASB, AS, and SAL developed the cytokine reservoir decay models. ASB, EL, AS, SoS, and SAL performed data visualization for the manuscript. SAL, ASB, EL, and SoS wrote the report with critical feedback the additional authors. Correspondence should be addressed to SAL.

## COMPETING INTERESTS STATEMENT

The authors declare no competing interests.

## Notes

### Competing Interest Statement

The authors have declared no competing interest.

### Author Declarations

The Human Research Protection Program/Institutional Review Board (IRB) at the University of California, San Francisco gave ethical approval for this work.

