## Supplementary Online Content for "NLRP3 Activation and Impaired TGF-β Anti-Inflammatory Pathways Predict Vascular Risk in PWH on ART"

**eMethods.** Quantification of plasma cytokines

**eTable1.** Baseline ART regimen of study participants

**eTable 2.** Differences in plasma soluble marker concentration detectability.

**eTable 3.** Cox proportional hazard model ASCVD risk estimates using both linear and nonlinear models.

**eTable 4.** Unadjusted and adjusted hazard ratios of incident vascular event (VE), which include ASCVD and VTE diagnoses.

**eTable 5.** Cox proportional hazard model ASCVD risk estimates of increased cytokine cluster medians using both linear and nonlinear models.

**eTable 6.** Direct effect estimates from doubling either IL-18 or TGF-β2.

**eTable 7.** Estimates of IL-18's direct and indirect effect on 5-year ASCVD risk.

**eFigure 1.** Study design and selection of participants and plasma samples

**eFigure 2.** Age, sex, race/ethnicity, and follow-up time of final selected case-cohort samples.

**eFigure 3.** Batch effects were detected in plate standards.

**eFigure 4.** Observed cytokine concentrations by plate before (A) and after (B) batch correction are shown for each cytokine.

**eFigure 5.** Cytokine concentrations of IL-18 and IP-10 across different vascular diagnoses.

**eFigure 6.** Comparison of plasma cytokine concentrations between any VE cases and controls.

**eFigure 7.** Hazard ratios of incident atherosclerotic cardiovascular disease (ASCVD).

**eFigure 8.** Unadjusted and adjusted hazard ratios of incident vascular event (VE), which include ASCVD and VTE diagnoses.

**eFigure 9.** Unadjusted and adjusted hazard ratios for increase in plasma cytokine cluster medians, as predictors of ASCVD risk.

**eFigure 10.** Higher levels of IL-18 but not TGF-β2 increase 5-year ASCVD risk in causal inference models.

**eFigure 11.** IL-18 as a persistent predictor of ASCVD risk after controlling for IL-6 and IL-1β.

### eMethods.

#### Quantification of plasma cytokines

Plasma concentrations of 33 cytokines and chemokines were measured, including fractalkine, I-TAC, IFN-γ, IL-10, IL-15, IL-18, IL-1β, IL-27, IL-29, IL-6, IL-7, IL-8, IL-9, IP-10, MCP-3, MIP-1α, MIP-3α, TGF-β1, TGF-β2, TNF-α, GM-CSF, IFN-α2a, IL-12p70, IL-17A, IL-2, IL-21, IL-22, IL-23, IL-33, IL-4, SDF-1α, and TGF-β3. Assays were performed on undiluted samples according to the manufacturer's protocol, and concentrations were calculated using MSD Discovery Workbench (version 4.0.13) with four-parameter logistic (4-PL) standard curves.

### eTable 1. Baseline ART regimen of study participants

| | Total (n=837) | PWH with ASCVD[a] (n=111) | PWH without ASCVD[a] (n=726) | *P* value | PWH with any VE[b] (n=135) | PWH without any VE[b] (n=702) | *P* value |
|---|---|---|---|---|---|---|---|
| **ART regimen, No. (%)** | | | | | | | |
| **Backbone drugs** | | | | | | | |
| **TDF-containing** | 346 (41.3) | 22 (19.8) | 324 (44.6) | <0.001 | 29 (21.5) | 317 (45.2) | <0.001 |
| **ABC-containing** | 91 (10.9) | 20 (18.0) | 71 (9.8) | 0.015 | 25 (18.5) | 66 (9.4) | 0.003 |
| **AZT-containing** | 248 (29.6) | 44 (39.6) | 204 (28.1) | 0.018 | 51 (37.8) | 197 (28.1) | 0.031 |
| **Other** | 216 (25.8) | 40 (36.0) | 176 (24.2) | 0.011 | 46 (34.1) | 170 (24.2) | 0.022 |
| **Non-Backbone Drugs** | | | | | | | |
| **INSTI-based** | 38 (4.5) | 4 (3.6) | 34 (4.7) | 0.792 | 33 (4.7) | 5 (3.7) | 0.776 |
| **NNRTI-based** | 366 (43.7) | 36 (32.4) | 330 (45.5) | 0.013 | 324 (46.2) | 42 (31.1) | 0.002 |
| **PI-based** | 408 (48.7) | 77 (69.4) | 331 (45.6) | <0.001 | 315 (44.9) | 93 (68.9) | <0.001 |
| **Other** | 114 (13.6) | 13 (11.7) | 101 (13.9) | 0.631 | 99 (14.1) | 15 (11.1) | 0.429 |

P values compare cases and controls; Mann–Whitney U tests were used for continuous variables and $\chi^2$ tests for categorical variables. TDF = tenofovir; ABC = abacavir INSTI, integrase strand transfer inhibitor; NNRTI, nonnucleoside reverse transcriptase inhibitor; PI, protease inhibitor.

**eTable 2. Differences in plasma soluble marker concentration detectability.** Limit of detection, percent detectable, percent quantifiable, and coefficient of variation are shown for each marker. The coefficient of variation is computed on samples above the detection limit. Results are sorted from highest to lowest percent quantifiable (above the detection limit).

| Soluble Marker | Limit of Detection[a] (pg/mL) | Percent Detectable[b] (%) | Percent Quantifiable[c] (%) | Coefficient of Variation[d] (%) |
|---|---|---|---|---|
| IL-15 | 0.82 | 98.72 | 98.72 | 36.13 |
| IP-10 | 0.49 | 98.72 | 98.72 | 89.92 |
| TGF-β1 | 9.10 | 98.82 | 98.62 | 53.95 |
| IL-8 | 0.15 | 98.72 | 98.22 | 1246.21 |
| TGF-β2 | 2.50 | 98.32 | 98.03 | 94.27 |
| IL-9 | 0.14 | 98.72 | 97.63 | 359.26 |
| IL-18 | 0.50 | 98.52 | 96.06 | 66.18 |
| TNF-α | 0.51 | 98.62 | 94.97 | 412.52 |
| IFN-γ | 1.70 | 96.84 | 94.77 | 272.47 |
| IL-7 | 1.50 | 98.72 | 93.49 | 53.63 |
| MIP-1α | 7.70 | 96.35 | 92.50 | 669.00 |
| MIP-3α | 1.80 | 94.67 | 92.31 | 189.82 |
| IL-27 | 9.60 | 97.04 | 91.62 | 212.78 |
| I-TAC | 1.50 | 96.94 | 85.70 | 145.23 |
| Fractalkine | 102.00 | 91.62 | 85.01 | 47.19 |
| SDF-1α | 280.00 | 83.63 | 80.08 | 64.57 |
| IL-6 | 0.33 | 95.86 | 79.49 | 1138.08 |
| IL-1β | 0.15 | 90.14 | 71.40 | 973.06 |
| IL-10 | 0.14 | 89.84 | 71.10 | 221.12 |
| IL-29/IFN-λ | 1.20 | 85.11 | 68.64 | 222.79 |
| MCP-3 | 0.79 | 97.04 | 59.57 | 278.76 |
| IL-33 | 0.59 | 72.58 | 54.34 | 182.98 |
| IL-4 | 0.08 | 64.69 | 54.04 | 384.55 |
| TGF-β3 | 1.40 | 84.02 | 49.80 | 101.64 |
| GM-CSF | 0.12 | 71.89 | 44.18 | 313.25 |
| IFN-β | 3.10 | 55.23 | 44.08 | 405.16 |
| IL-17A | 2.10 | 68.93 | 33.83 | 351.84 |
| IL-21 | 1.20 | 38.46 | 33.04 | 881.57 |
| IL-22 | 0.13 | 34.91 | 27.91 | 987.43 |
| IL-23 | 1.40 | 16.07 | 15.19 | 45.95 |
| IL-12p70 | 0.69 | 80.97 | 15.09 | 315.23 |
| IFN-α2a | 4.00 | 80.08 | 6.31 | 87.75 |
| IL-2 | 0.70 | 38.07 | 4.54 | 144.75 |

[a] Limit of detection = assay marker-specific limit of detection.
[b] Percent detectable = % samples that were non-zero.
[c] Percent quantifiable = % of samples above the limit of detection.
[d] Coefficient of Variation % = 100 x (standard deviation / mean)

**eTable 3. Cox proportional hazard model ASCVD risk estimates using both linear and nonlinear models.** Hazard ratios (HRs) and confidence intervals using robust standard errors from the linear models are reported per 2-fold increase in plasma cytokine concentrations. The nonlinear Cox model reports the significance of a degree-4 cubic B-spline fit to cytokine concentrations on a log2 scale. Separate models were fit for each cytokine, with both linear and nonlinear approaches, and for each model type (unadjusted and adjusted). False discovery rate (FDR) corrected p-values (q-values) are provided for each model.

### A. Linear Cox PH Models

| | | Unadjusted Cox PH Model | | | | Adjusted Cox PH Model | | | |
|---|---|---|---|---|---|---|---|---|---|
| **Cytokine** | **N** | **HR** | **95% CI** | **p-value** | **q-value** | **aHR[a]** | **95% CI** | **p-value** | **q-value** |
| IL-15 | 824 | 1.92 | (1.08, 3.43) | 0.026 | 0.106 | 1.68 | (0.86, 3.27) | 0.129 | 0.368 |
| IL-18 | 822 | 1.89 | (1.33, 2.68) | 0.000 | 0.007 | 1.57 | (1.07, 2.30) | 0.021 | 0.368 |
| Fractalkine | 801 | 1.67 | (0.98, 2.82) | 0.058 | 0.129 | 1.33 | (0.76, 2.32) | 0.312 | 0.567 |
| TNF-α | 824 | 1.43 | (1.06, 1.93) | 0.020 | 0.102 | 1.31 | (0.94, 1.83) | 0.115 | 0.368 |
| IP-10 | 824 | 1.38 | (1.05, 1.81) | 0.020 | 0.102 | 1.32 | (0.96, 1.82) | 0.084 | 0.368 |
| IL-7 | 824 | 1.31 | (0.89, 1.92) | 0.167 | 0.279 | 1.25 | (0.82, 1.91) | 0.291 | 0.567 |
| IL-8 | 824 | 1.28 | (0.95, 1.72) | 0.108 | 0.216 | 1.08 | (0.76, 1.54) | 0.665 | 0.782 |
| IL-6 | 797 | 1.20 | (1.00, 1.44) | 0.056 | 0.129 | 1.03 | (0.81, 1.30) | 0.815 | 0.847 |
| IL-27 | 821 | 1.18 | (0.92, 1.50) | 0.193 | 0.297 | 1.08 | (0.81, 1.43) | 0.613 | 0.766 |
| IFN-γ | 821 | 1.17 | (1.00, 1.36) | 0.054 | 0.129 | 1.16 | (0.97, 1.38) | 0.107 | 0.368 |
| MCP-3 | 814 | 1.10 | (0.92, 1.33) | 0.301 | 0.430 | 1.08 | (0.88, 1.33) | 0.434 | 0.619 |
| IFN-λ | 707 | 1.10 | (0.96, 1.26) | 0.164 | 0.279 | 1.12 | (0.97, 1.29) | 0.112 | 0.368 |
| I-TAC | 817 | 1.09 | (0.85, 1.41) | 0.494 | 0.618 | 1.19 | (0.89, 1.58) | 0.246 | 0.548 |
| MIP-3α | 807 | 1.06 | (0.93, 1.22) | 0.370 | 0.494 | 1.05 | (0.91, 1.20) | 0.539 | 0.719 |
| MIP-1α | 818 | 1.02 | (0.86, 1.20) | 0.826 | 0.918 | 0.98 | (0.78, 1.22) | 0.847 | 0.847 |
| IL-1β | 784 | 0.99 | (0.84, 1.17) | 0.921 | 0.955 | 1.03 | (0.87, 1.21) | 0.770 | 0.847 |
| IL-9 | 824 | 0.99 | (0.72, 1.36) | 0.955 | 0.955 | 0.86 | (0.61, 1.21) | 0.389 | 0.599 |
| IL-10 | 782 | 0.97 | (0.84, 1.12) | 0.651 | 0.766 | 1.08 | (0.92, 1.28) | 0.352 | 0.587 |
| TGF-β1 | 816 | 0.85 | (0.72, 1.00) | 0.055 | 0.129 | 0.90 | (0.76, 1.06) | 0.196 | 0.491 |

| | | | | | | | | | |
|---|---|---|---|---|---|---|---|---|---|
| TGF-β2 | 815 | 0.74 | (0.61, 0.90) | 0.003 | 0.026 | 0.84 | (0.67, 1.05) | 0.128 | 0.368 |

[a]Adjusted hazard ratio (aHR) is adjusted for traditional vascular risk factors – age at plasma sample, sex, race/ethnicity, hyperlipidemia, hypertension, type II diabetes, smoking status, and family history of vascular disease

### B. Nonlinear Cox PH Models

| | | Unadjusted Cox PH Model | | | Adjusted Cox PH Model | | |
|---|---|---|---|---|---|---|---|
| **Cytokine** | **N** | **Wald Statistic (df=4)** | **p-value** | **q-value** | **aWald Statistic[a] (df=4)** | **p-value** | **q-value** |
| IL-15 | 824 | 9.21 | 0.056 | 0.187 | 7.44 | 0.114 | 0.400 |
| IL-18 | 822 | 17.34 | 0.002 | 0.033 | 8.66 | 0.070 | 0.400 |
| Fractalkine | 801 | 10.62 | 0.031 | 0.125 | 4.21 | 0.378 | 0.504 |
| TNF-α | 824 | 5.42 | 0.246 | 0.352 | 5.14 | 0.273 | 0.498 |
| IP-10 | 824 | 6.06 | 0.195 | 0.324 | 3.49 | 0.480 | 0.565 |
| IL-7 | 824 | 7.47 | 0.113 | 0.229 | 5.73 | 0.220 | 0.498 |
| IL-8 | 824 | 6.19 | 0.186 | 0.324 | 2.45 | 0.653 | 0.688 |
| IL-6 | 797 | 11.78 | 0.019 | 0.095 | 4.89 | 0.299 | 0.498 |
| IL-27 | 821 | 7.43 | 0.115 | 0.229 | 2.67 | 0.615 | 0.683 |
| IFN-γ | 821 | 8.72 | 0.069 | 0.196 | 7.32 | 0.120 | 0.400 |
| MCP-3 | 814 | 8.04 | 0.090 | 0.225 | 4.65 | 0.325 | 0.500 |
| IFN-λ | 707 | 3.73 | 0.443 | 0.529 | 4.30 | 0.366 | 0.504 |
| I-TAC | 817 | 1.84 | 0.765 | 0.765 | 5.02 | 0.285 | 0.498 |
| MIP-3α | 807 | 5.13 | 0.275 | 0.366 | 1.80 | 0.772 | 0.772 |
| MIP-1α | 818 | 12.81 | 0.012 | 0.082 | 10.63 | 0.031 | 0.400 |
| IL-1β | 784 | 5.76 | 0.218 | 0.336 | 7.33 | 0.120 | 0.400 |
| IL-9 | 824 | 2.68 | 0.612 | 0.680 | 3.93 | 0.415 | 0.519 |
| IL-10 | 782 | 2.24 | 0.691 | 0.728 | 5.67 | 0.225 | 0.498 |
| TGF-β1 | 816 | 3.69 | 0.450 | 0.529 | 6.06 | 0.195 | 0.498 |
| TGF-β2 | 815 | 14.14 | 0.007 | 0.069 | 8.58 | 0.073 | 0.400 |

[a]Adjusted Wald statistic (aWald) is adjusted for age at plasma sample, sex, race/ethnicity, hyperlipidemia, hypertension, type II diabetes, smoking status, and family history of vascular disease

**eTable 4. Unadjusted and adjusted hazard ratios of incident vascular event (VE), which include ASCVD and VTE diagnoses.** Hazard ratios (HRs) and confidence intervals using robust standard errors from the linear models are reported per 2-fold increase in plasma cytokine concentrations. The nonlinear Cox model reports the significance of a degree-4 cubic B-spline fit to cytokine concentrations on a log2 scale. Separate models were fit for each cytokine, with both linear and nonlinear approaches, and for each model type (unadjusted and adjusted). False discovery rate (FDR) corrected p-values (q-values) are provided for each model.

### A. Linear Cox PH Models

| | | Unadjusted Cox PH Model | | | | Adjusted Cox PH Model | | | |
|---|---|---|---|---|---|---|---|---|---|
| **Cytokine** | **N** | **HR** | **95% CI** | **p-value** | **q-value** | **aHR[a]** | **95% CI** | **p-value** | **q-value** |
| IL-15 | 837 | 1.81 | (1.06, 3.08) | 0.029 | 0.193 | 1.56 | (0.86, 2.84) | 0.148 | 0.826 |
| IL-18 | 835 | 1.67 | (1.21, 2.29) | 0.002 | 0.035 | 1.37 | (0.98, 1.93) | 0.068 | 0.826 |
| Fractalkine | 814 | 1.52 | (0.92, 2.51) | 0.100 | 0.373 | 1.18 | (0.70, 1.98) | 0.538 | 0.826 |
| TNF-α | 837 | 1.20 | (0.90, 1.61) | 0.213 | 0.473 | 1.10 | (0.80, 1.51) | 0.562 | 0.826 |
| IP-10 | 837 | 1.24 | (0.95, 1.61) | 0.115 | 0.373 | 1.16 | (0.86, 1.57) | 0.338 | 0.826 |
| IL-7 | 837 | 1.17 | (0.82, 1.68) | 0.378 | 0.612 | 1.15 | (0.79, 1.68) | 0.476 | 0.826 |
| IL-8 | 837 | 1.12 | (0.84, 1.49) | 0.428 | 0.612 | 0.97 | (0.70, 1.33) | 0.844 | 0.899 |
| IL-6 | 810 | 1.13 | (0.95, 1.34) | 0.162 | 0.405 | 0.96 | (0.78, 1.19) | 0.717 | 0.843 |
| IL-27 | 834 | 1.10 | (0.87, 1.38) | 0.423 | 0.612 | 1.02 | (0.78, 1.32) | 0.899 | 0.899 |
| IFN-γ | 833 | 1.12 | (0.97, 1.29) | 0.131 | 0.373 | 1.11 | (0.94, 1.30) | 0.215 | 0.826 |
| MCP-3 | 827 | 1.08 | (0.91, 1.27) | 0.381 | 0.612 | 1.05 | (0.88, 1.26) | 0.585 | 0.826 |
| IFN-λ | 718 | 1.07 | (0.95, 1.21) | 0.260 | 0.520 | 1.10 | (0.97, 1.24) | 0.124 | 0.826 |
| I-TAC | 830 | 1.02 | (0.79, 1.30) | 0.904 | 0.904 | 1.07 | (0.81, 1.41) | 0.628 | 0.826 |
| MIP-3α | 820 | 1.03 | (0.91, 1.17) | 0.633 | 0.791 | 1.01 | (0.89, 1.15) | 0.858 | 0.899 |
| MIP-1α | 831 | 0.99 | (0.83, 1.18) | 0.889 | 0.904 | 0.94 | (0.74, 1.18) | 0.580 | 0.826 |
| IL-1β | 796 | 1.01 | (0.87, 1.18) | 0.899 | 0.904 | 1.04 | (0.89, 1.20) | 0.633 | 0.826 |
| IL-9 | 837 | 1.04 | (0.78, 1.38) | 0.805 | 0.904 | 0.91 | (0.67, 1.24) | 0.547 | 0.826 |
| IL-10 | 795 | 0.95 | (0.83, 1.09) | 0.474 | 0.631 | 1.04 | (0.89, 1.21) | 0.661 | 0.826 |

| | | | | | | | | | |
|---|---|---|---|---|---|---|---|---|---|
| TGF-β1 | 829 | 0.88 | (0.75, 1.03) | 0.104 | 0.373 | 0.92 | (0.78, 1.09) | 0.345 | 0.826 |
| TGF-β2 | 827 | 0.80 | (0.67, 0.95) | 0.013 | 0.125 | 0.92 | (0.75, 1.13) | 0.416 | 0.826 |

[a]Adjusted hazard ratio (aHR) is adjusted for traditional vascular risk factors – age at plasma sample, sex, race/ethnicity, hyperlipidemia, hypertension, type II diabetes, smoking status, and family history of vascular disease

### B. Nonlinear Cox PH Models

| | | Unadjusted Cox PH Model | | | Adjusted Cox PH Model | | |
|---|---|---|---|---|---|---|---|
| **Cytokine** | **N** | **Wald Statistic (df=4)** | **p-value** | **q-value** | **aWald Statistic[a] (df=4)** | **p-value** | **q-value** |
| IL-15 | 837 | 10.47 | 0.033 | 0.133 | 7.61 | 0.107 | 0.616 |
| IL-18 | 835 | 12.55 | 0.014 | 0.130 | 5.48 | 0.242 | 0.622 |
| Fractalkine | 814 | 11.05 | 0.026 | 0.130 | 4.26 | 0.372 | 0.622 |
| TNF-α | 837 | 2.07 | 0.724 | 0.762 | 3.57 | 0.467 | 0.622 |
| IP-10 | 837 | 4.00 | 0.406 | 0.588 | 3.28 | 0.512 | 0.640 |
| IL-7 | 837 | 5.55 | 0.235 | 0.475 | 5.15 | 0.272 | 0.622 |
| IL-8 | 837 | 3.75 | 0.441 | 0.588 | 2.86 | 0.582 | 0.666 |
| IL-6 | 810 | 11.50 | 0.022 | 0.130 | 4.44 | 0.350 | 0.622 |
| IL-27 | 834 | 6.79 | 0.147 | 0.368 | 2.38 | 0.666 | 0.666 |
| IFN-γ | 833 | 8.40 | 0.078 | 0.260 | 6.82 | 0.146 | 0.616 |
| MCP-3 | 827 | 7.76 | 0.101 | 0.288 | 3.88 | 0.422 | 0.622 |
| IFN-λ | 718 | 2.63 | 0.621 | 0.690 | 4.07 | 0.396 | 0.622 |
| I-TAC | 830 | 1.34 | 0.854 | 0.854 | 3.95 | 0.413 | 0.622 |
| MIP-3α | 820 | 5.52 | 0.238 | 0.475 | 2.51 | 0.642 | 0.666 |
| MIP-1α | 831 | 5.26 | 0.262 | 0.476 | 2.48 | 0.648 | 0.666 |
| IL-1β | 796 | 4.70 | 0.320 | 0.533 | 6.68 | 0.154 | 0.616 |
| IL-9 | 837 | 3.75 | 0.441 | 0.588 | 5.97 | 0.202 | 0.622 |
| IL-10 | 795 | 2.79 | 0.593 | 0.690 | 6.86 | 0.143 | 0.616 |
| TGF-β1 | 829 | 2.67 | 0.615 | 0.690 | 3.69 | 0.449 | 0.622 |
| TGF-β2 | 827 | 12.96 | 0.011 | 0.130 | 8.29 | 0.082 | 0.616 |

[a]Adjusted Wald statistic (aWald) is adjusted for age at plasma sample, sex, race/ethnicity, hyperlipidemia, hypertension, type II diabetes, smoking status, and family history of vascular disease

**eTable 5. Cox proportional hazard model ASCVD risk estimates of increased cytokine cluster medians using both linear and nonlinear models.** Hazard ratios (HRs) and confidence intervals using robust standard errors from the linear models are reported per one unit increase in the median value of the cytokine cluster after log-transforming and scaling concentrations of each cytokine in the cluster to mean zero and unit variance. The nonlinear Cox model reports the significance of a degree-4 cubic B-spline fit to cytokine concentrations on a log2 scale. Separate models were fit for each cytokine, with both linear and nonlinear approaches, and for each model type (unadjusted and adjusted). False discovery rate (FDR) corrected p-values (q-values) are provided for each model.

### A. Linear Cox PH Models

| | | Unadjusted Cox PH Model | | | | Adjusted Cox PH Model | | | |
|---|---|---|---|---|---|---|---|---|---|
| **Cytokine Cluster** | **N** | **HR** | **95% CI** | **p-value** | **q-value** | **aHR[a]** | **95% CI** | **p-value** | **q-value** |
| Cluster 1: Fractalkine | 824 | 1.20 | (0.99, 1.44) | 0.058 | 0.155 | 1.11 | (0.91, 1.34) | 0.312 | 0.519 |
| Cluster 2: I-TAC, IL-27, MIP-3α | 824 | 1.22 | (0.95, 1.55) | 0.115 | 0.225 | 1.14 | (0.88, 1.48) | 0.326 | 0.519 |
| Cluster 3: IFN-γ, IL-15, IP-10, MCP-3, MIP-1α, TNF-α | 824 | 1.35 | (1.07, 1.69) | 0.011 | 0.045 | 1.26 | (0.97, 1.63) | 0.082 | 0.384 |
| Cluster 4: IL-10, IFN-λ | 824 | 1.01 | (0.82, 1.25) | 0.916 | 0.955 | 1.19 | (0.93, 1.51) | 0.160 | 0.427 |
| Cluster 5: IL-18, IL-6 | 824 | 1.53 | (1.21, 1.94) | 0.000 | 0.003 | 1.27 | (0.96, 1.69) | 0.096 | 0.384 |
| Cluster 6: IL-1β, IL-8 | 824 | 1.12 | (0.91, 1.39) | 0.296 | 0.394 | 1.07 | (0.86, 1.34) | 0.556 | 0.573 |
| Cluster 7: IL-7, TGF-β1, TGF-β2 | 824 | 0.83 | (0.64, 1.07) | 0.140 | 0.225 | 0.93 | (0.71, 1.21) | 0.573 | 0.573 |
| Cluster 8: IL-9 | 824 | 0.99 | (0.79, 1.25) | 0.955 | 0.955 | 0.90 | (0.70, 1.15) | 0.389 | 0.519 |

[a]Adjusted hazard ratio (aHR) is adjusted for traditional vascular risk factors – age at plasma sample, sex, race/ethnicity, hyperlipidemia, hypertension, type II diabetes, smoking status, and family history of vascular disease

### B. Nonlinear Cox PH Models

| | | Unadjusted Cox PH Model | | | Adjusted Cox PH Model | | |
|---|---|---|---|---|---|---|---|
| **Cytokine Cluster** | **N** | **Wald Statistic (df=4)** | **p-value** | **q-value** | **aWald Statistic[a] (df=4)** | **p-value** | **q-value** |
| Cluster 1: Fractalkine | 824 | 10.62 | 0.031 | 0.083 | 4.21 | 0.378 | 0.474 |
| Cluster 2: I-TAC, IL-27, MIP-3α | 824 | 7.48 | 0.113 | 0.180 | 5.23 | 0.265 | 0.474 |
| Cluster 3: IFN-γ, IL-15, IP-10, MCP-3, MIP-1α, TNF-α | 824 | 10.71 | 0.030 | 0.083 | 8.56 | 0.073 | 0.474 |

| | | | | | | | |
|---|---|---|---|---|---|---|---|
| Cluster 4: IL-10, IFN-λ | 824 | 2.07 | 0.723 | 0.723 | 4.71 | 0.318 | 0.474 |
| Cluster 5: IL-18, IL-6 | 824 | 17.27 | 0.002 | 0.014 | 6.65 | 0.155 | 0.474 |
| Cluster 6: IL-1β, IL-8 | 824 | 4.48 | 0.345 | 0.460 | 1.90 | 0.755 | 0.755 |
| Cluster 7: IL-7, TGF-β1, TGF-β2 | 824 | 8.10 | 0.088 | 0.176 | 6.23 | 0.182 | 0.474 |
| Cluster 8: IL-9 | 824 | 2.68 | 0.612 | 0.700 | 3.93 | 0.415 | 0.474 |

[a]Adjusted Wald statistic (aWald) is adjusted for age at plasma sample, sex, race/ethnicity, hyperlipidemia, hypertension, type II diabetes, smoking status, and family history of vascular disease

**eTable 6. Direct effect estimates from doubling either IL-18 or TGF-β2.** Absolute risk and relative risk increase estimates are provided from causal models with standard errors (SE), 95% CI, and p-values. All models are adjusted for traditional ASCVD risk factors (age, sex, race/ethnicity, family history of vascular disease, tobacco use, comorbidities (diagnosis of hyperlipidemia, hypertension, or type II diabetes), and HIV clinical factors (timing of ART initiation, duration of viral suppression, nadir CD4+ T count, pre-ART viral load, and ART regimen). A secondary model of the effect of IL-18 on 5-year ASCVD risk also adjusted for IL-1β because of its membership in the NLRP3 inflammasome.

| Cytokine | Absolute Risk | SE | 95% C.I. | p-value | Relative Risk | SE | 95% CI | p-value |
|---|---|---|---|---|---|---|---|---|
| **IL-18** | 0.0363 | 0.0416 | (-0.045, 0.118) | 0.383 | 2.12 | 0.5913 | (0.667, 6.768) | 0.203 |
| **IL-18**[a] | 0.0415 | 0.0386 | (-0.034, 0.117) | 0.282 | 2.31 | 0.5135 | (0.845, 6.326) | 0.103 |
| **TGF-β2** | 0.0073 | 0.0064 | (-0.005, 0.020) | 0.251 | 1.22 | 0.1627 | (0.890, 1.684) | 0.214 |

[a] denotes that the model additionally included IL-1β as a possible confounder.

**eTable 7. Estimates of IL-18's direct and indirect effect on 5-year ASCVD risk.** Absolute risk increase estimates are provided from causal models along with standard errors, 95% CI, and p-values. All models are adjusted for traditional ASCVD risk factors (age, sex, race/ethnicity, family history of vascular disease, tobacco use, comorbidities (diagnosis of hyperlipidemia, hypertension, or type II diabetes), and HIV clinical factors (timing of ART initiation, duration of viral suppression, nadir CD4+ T count, pre-ART viral load, and ART regimen). A secondary model also adjusted for IL-1β because of its membership in the NLRP3 inflammasome.

**A. Adjusted for ASCVD and HIV factors**

| **Pathway** | **Risk Increase** | **Standard error** | **95% CI** | **p-value** |
|---|---|---|---|---|
| **IL-18 → ASCVD** | 0.0149 | 0.00599 | (0.00313, 0.0266) | 1.30e-2 |
| **IL-18 → IL-6 → ASCVD** | 0.0428 | 0.00337 | (0.0362, 0.0494) | 5.02e-37 |
| **Total Effect of IL-18** | 0.0577 | 0.00558 | (0.0468, 0.0686) | 4.36e-25 |

**B. Adjusted for IL-1β in addition to ASCVD and HIV factors**

| **Pathway** | **Risk Increase** | **Standard error** | **95% CI** | **p-value** |
|---|---|---|---|---|
| **IL-18 → ASCVD** | 0.0260 | 0.00533 | (0.0155, 0.0364) | 1.09e-6 |
| **IL-18 → IL-6 → ASCVD** | 0.0734 | 0.00420 | (0.0652, 0.0816) | 1.96e-68 |
| **Total Effect of IL-18** | 0.0994 | 0.00647 | (0.0867, 0.112) | 2.94e-53 |

**eFigure 1. Study design and sample selection overview of plasma cytokines associated with incident vascular events (VE) in people with HIV (PWH) on antiretroviral therapy (ART).** A total of 2,239 PWH from the U.S. Military HIV Natural History Study with biospecimen availability during ART suppression (at least one year of ART, sustained through follow-up) were eligible for inclusion. VE cases were defined as incident diagnoses of atherosclerotic cardiovascular disease (ASCVD) – coronary artery disease (CAD), myocardial infarction (MI), stroke (CVA), peripheral artery disease (PAD) – as well as diagnoses of venous thrombotic events (VTE) – deep vein thrombosis (DVT) and pulmonary embolism (PE). We used a stratified case-cohort design to select 1,002 participants. To avoid potential reverse causation (VE causing changes in cytokine levels) and effects of non-suppressed HIV, we excluded participants with prior VE or who were not virally suppressed for at least one year at the time of plasma sample.

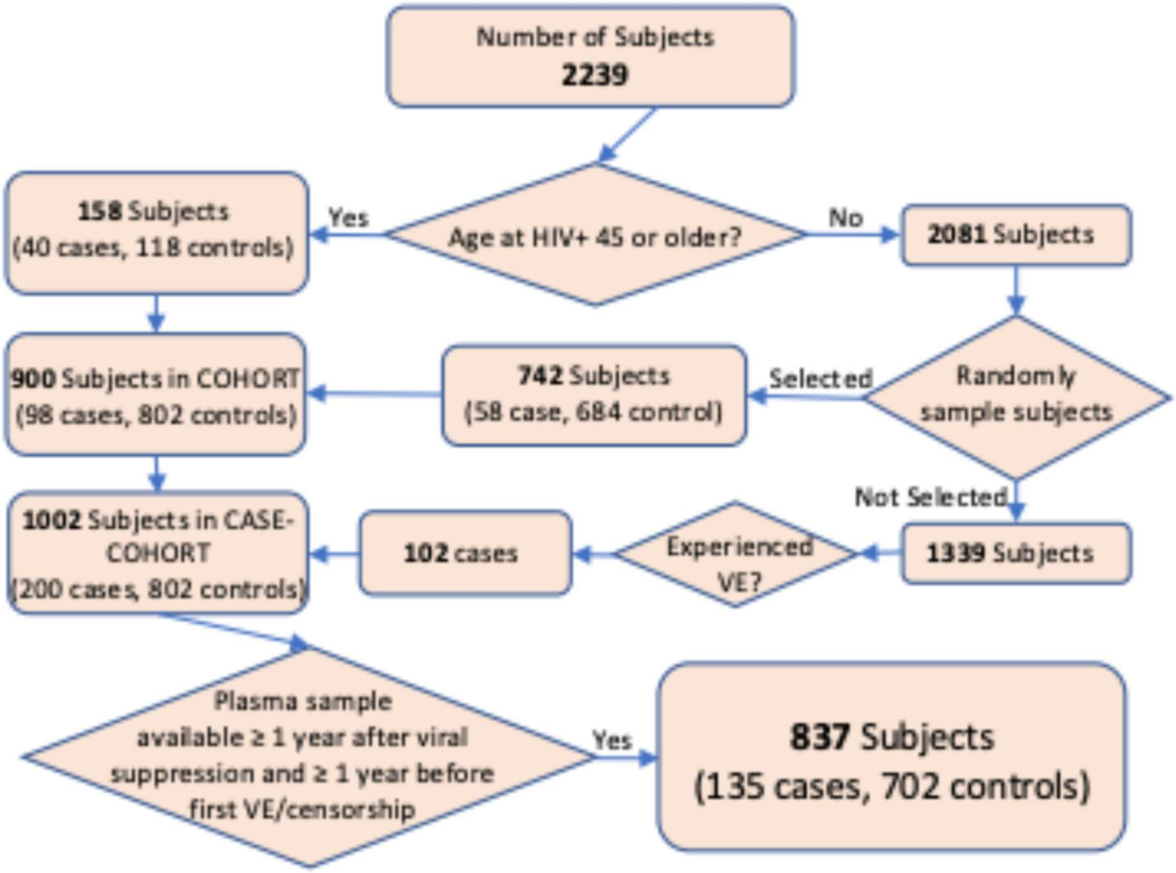

**eFigure 2. Age, sex, race/ethnicity, and follow-up time of final selected case-cohort samples.** We used a stratified case-cohort design to select 1,002 participants. Since age and male sex are established risk factors for vascular disease, we oversampled controls to achieve comparable age and sex distributions across groups. Results of sample selection show relatively comparable age (at plasma sample) (A), follow-up time (B), sex (C), and race/ethnicity (D) distributions by group. ASCVD = atherosclerotic cardiovascular disease; any new onset coronary artery disease (CAD), myocardial infarction (MI), cerebrovascular accident (CVA), or peripheral arterial disease (PAD) diagnosis. Venous thrombotic event (VTE); any new onset deep vein thrombosis (DVT) or pulmonary embolism (PE). VE = vascular event; composite outcome of any incident ASCVD or VTE.

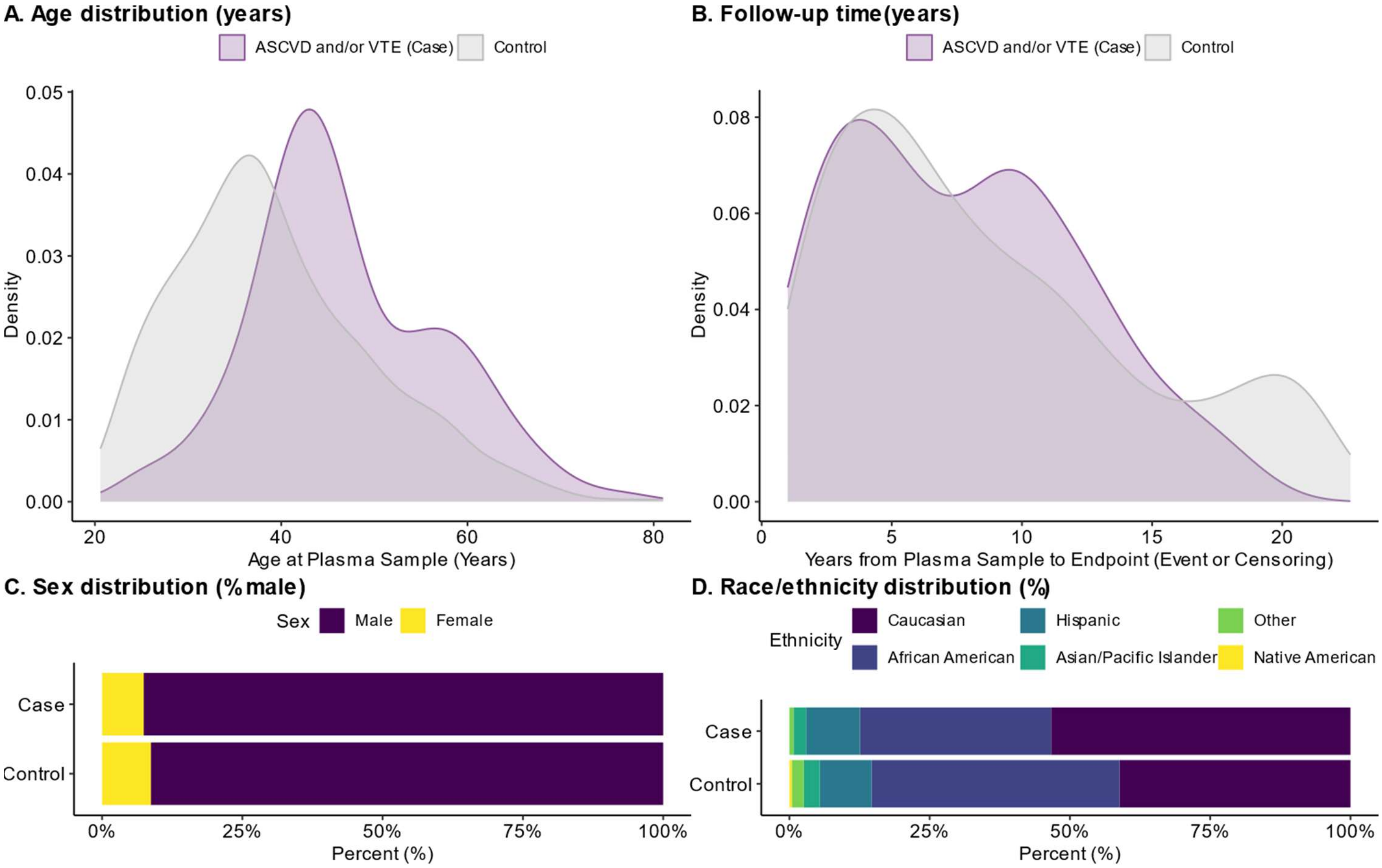

**eFigure 3. Batch effects were detected in plate standards.** Comparison of standards (of known quantities for each cytokine) were run for each of the 29 plates. A subset of the plates demonstrated evidence of batch effects (more than two of the standard measurements were more than two median absolute deviations from the median standard value) as shown in black ("Outlier Plates") compared to green ("Typical Plates") lines for each cytokine. These results suggest the need for batch-correction to ensure that cytokine measurements could be compared across plates in subsequent analyses.

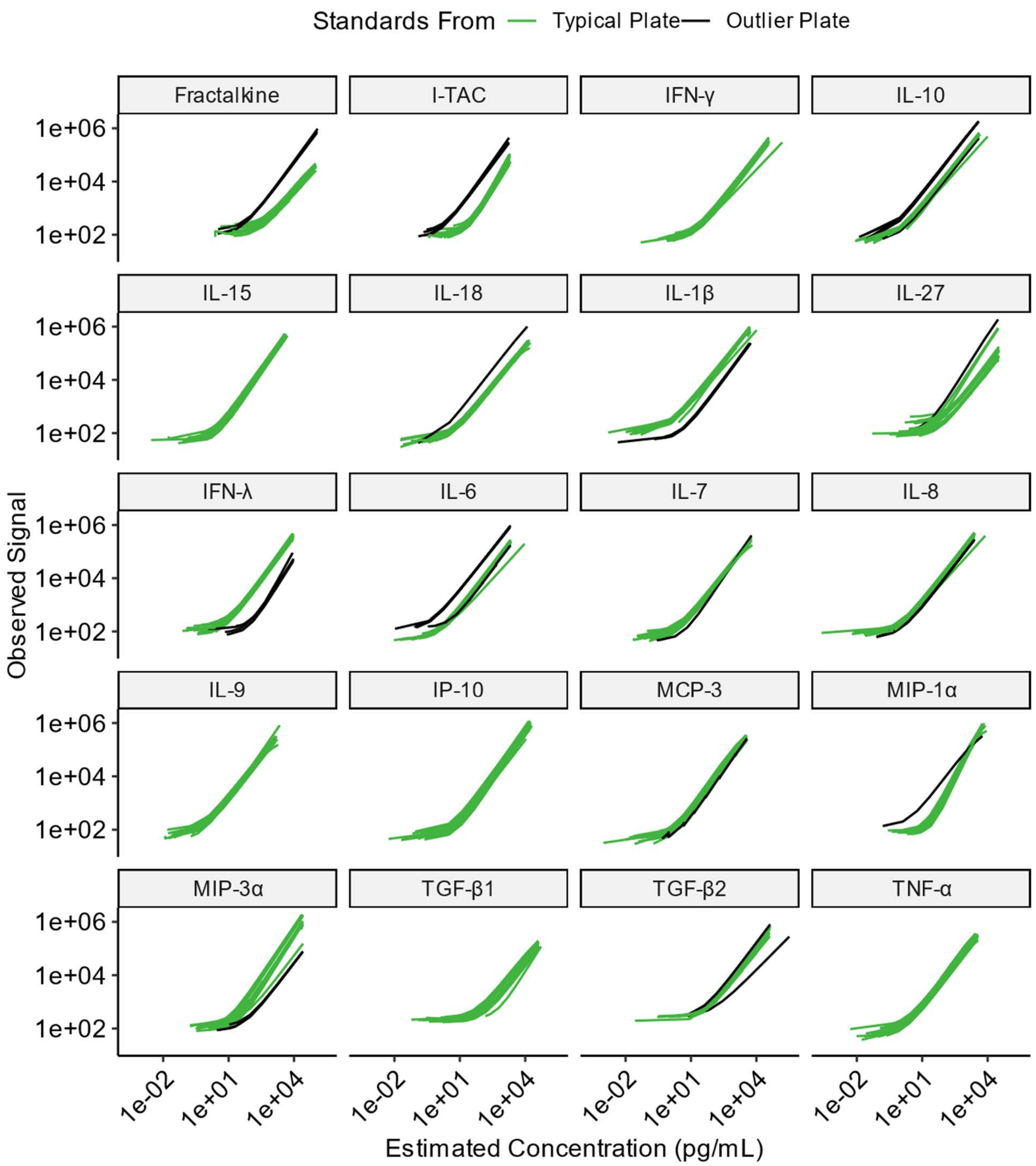

**eFigure 4. Observed cytokine concentrations by plate before (A) and after (B) batch correction are shown for each cytokine.** Density plots demonstrate observed data in black (“Outlier Plate”) and green (“Typical Plate”) for each cytokine. After batch correction, the black and green density plots are now more closely aligned.

**A.**

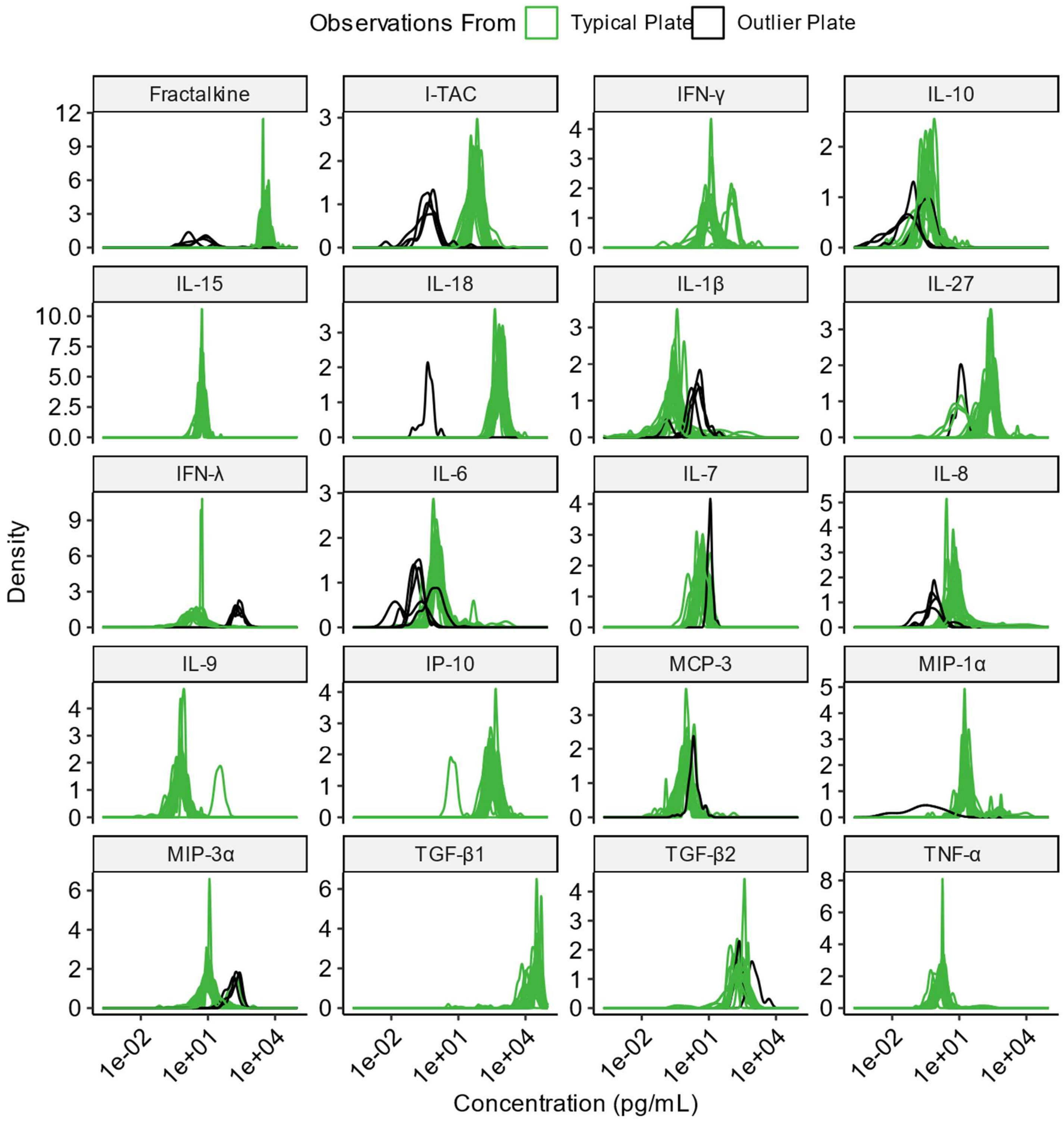

**B.**

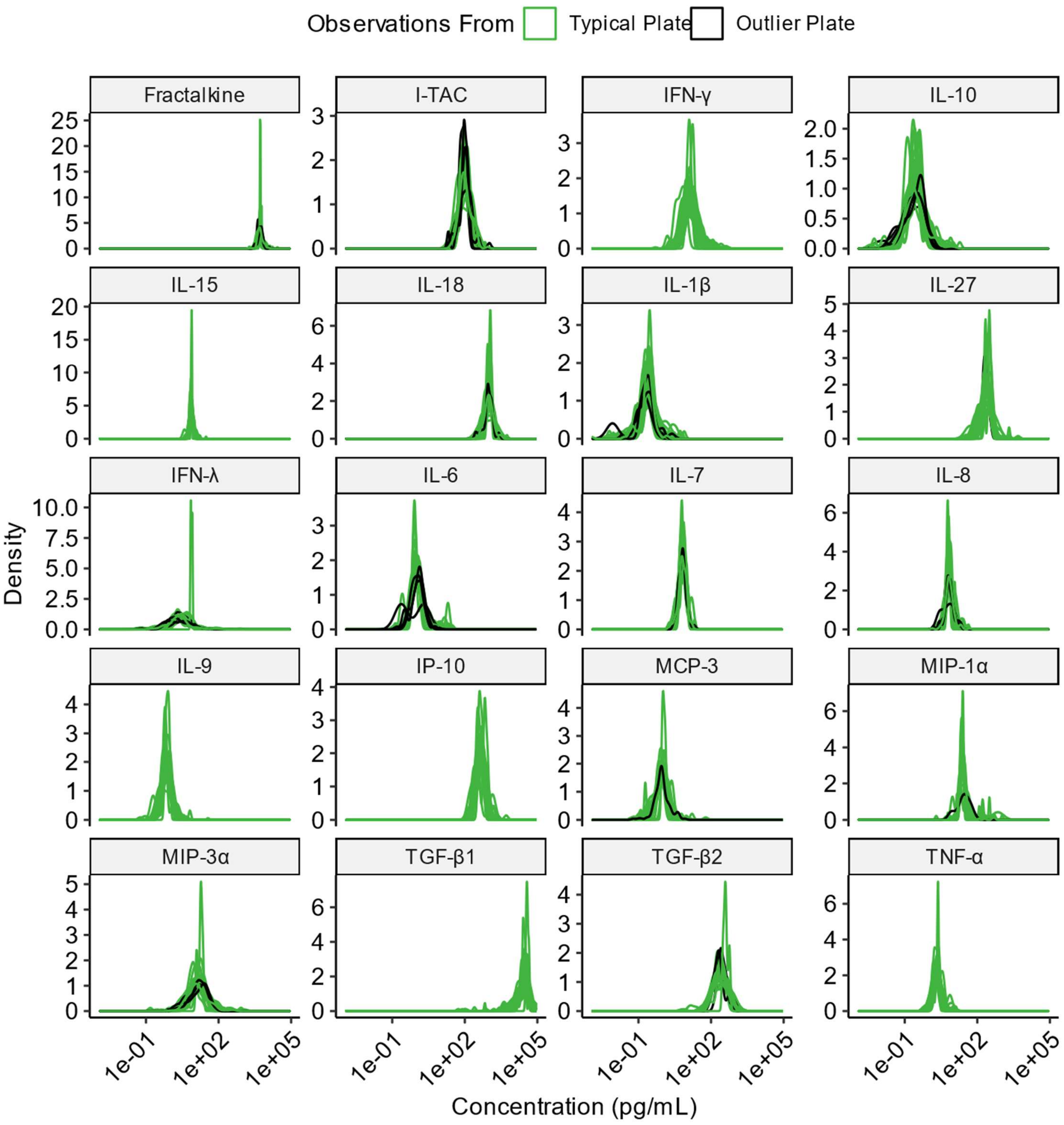

Observations From
Typical Plate
Outlier Plate
Fractalkine
I-TAC
IFN-γ
IL-10
IL-15
IL-18
IL-1β
IL-27
IFN-λ
IL-6
IL-7
IL-8
IL-9
IP-10
MCP-3
MIP-1α
MIP-3α
TGF-β1
TGF-β2
TNF-α
Density
1e-01
1e+02
1e+05
Concentration (pg/mL)

**eFigure 5. Cytokine concentrations of IL-18 and IP-10 across different vascular diagnoses.** Disaggregated Mann-Whitney tests for IL-18 (**A**) and IP-10 (**B**) concentrations are shown by specific incident VE diagnoses. IL-18 was elevated in participants with each type of ASCVD (red boxes) compared to controls. In contrast, IP-10 levels were only elevated in participants with coronary artery disease (CAD) and myocardial infarction (MI). Neither cytokine showed significant differences in the two types of venous thrombotic event (VTE) outcomes (blue boxes), though levels appeared to be slightly lower in these groups.

**A.**

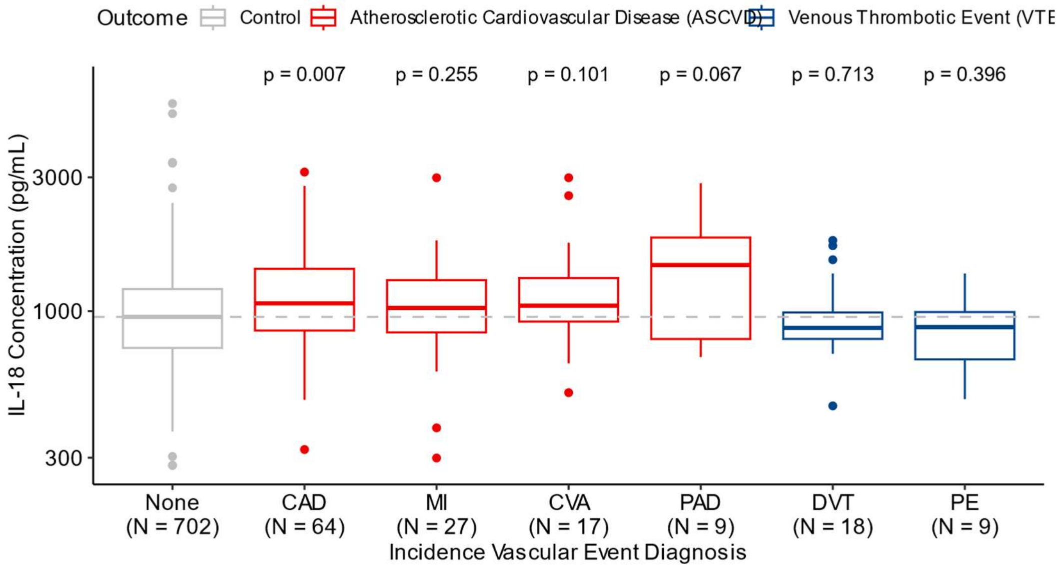


**B.**

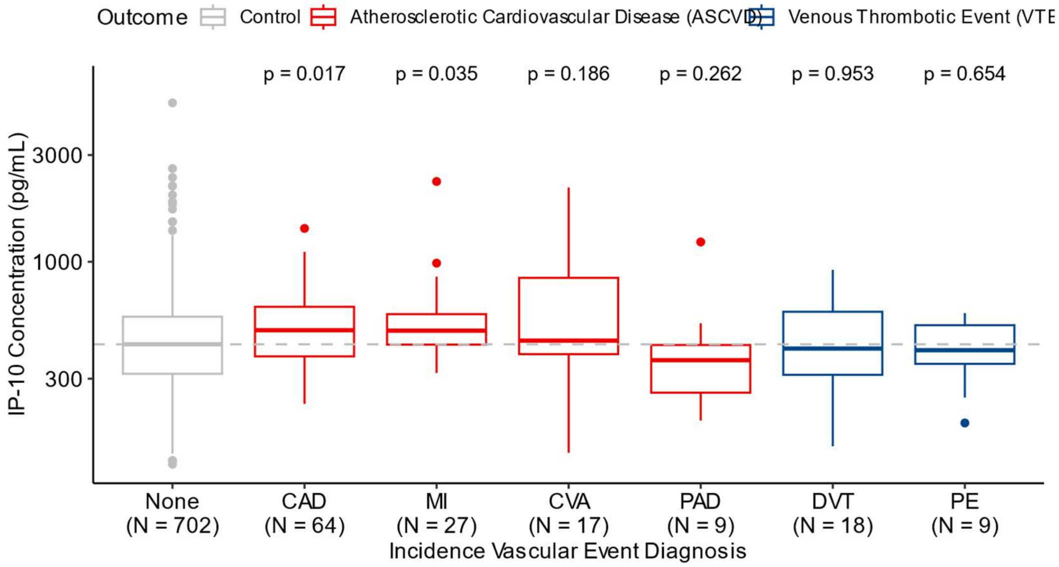


*Abbreviations*: CAD: Coronary artery disease; MI: Myocardial infarction; CVA: Cerebrovascular accident (including ischemic and hematologic stroke, vertebrobasilar insufficiency, etc.); PAD: Peripheral artery disease; DVT: Deep vein thrombosis; PE: Pulmonary embolism.

**eFigure 6. Comparison of plasma cytokine concentrations between any VE cases and controls.** This figure compares plasma cytokine concentrations between any VE (ASCVD and/or VTE) cases and controls using Mann-Whitney U tests. Significance levels are indicated as follows: *** for $p < 0.001$, ** for $p < 0.01$, * for $p < 0.05$, and ‘ns’ for $p \geq 0.05$. All statistically significant cytokines ($p < 0.05$) were elevated in VE cases relative to controls; however, no cytokines were found to be significant after FDR correction.

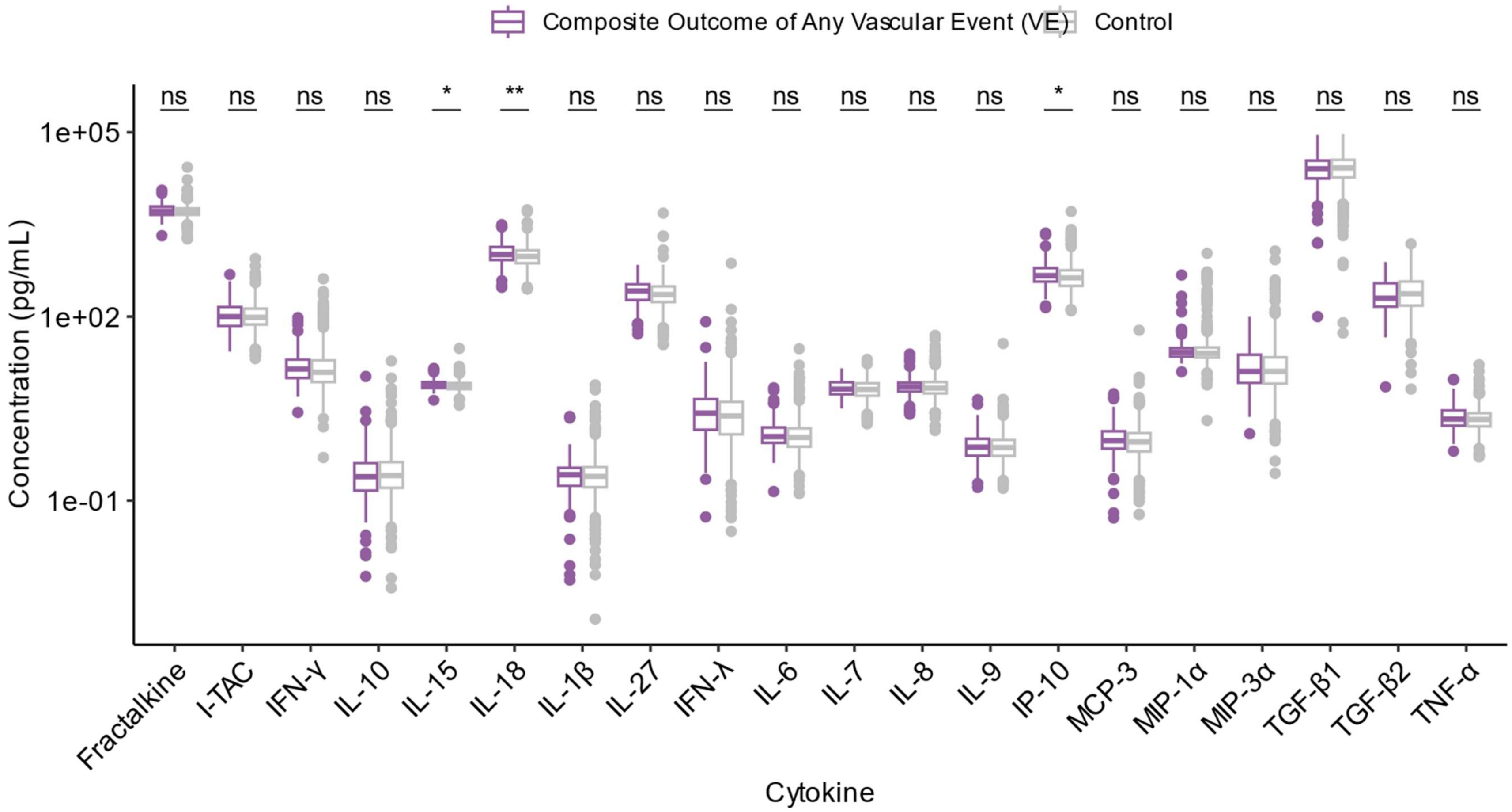

**eFigure 7. Hazard ratios of incident atherosclerotic cardiovascular disease (ASCVD).** We fit multivariate linear Cox proportional hazards models to determine whether inclusion of traditional vascular risk factors or HIV clinical factors in our models altered our risk estimates. Since age is a strong predictor of ASCVD (and a traditional vascular risk factor as well an important HIV clinical factor), all adjusted models included age (at plasma sample) as a covariate. Traditional vascular risk factors included sex, race/ethnicity, family history of vascular disease, tobacco use, and comorbidities (diagnosis of hyperlipidemia, hypertension, or type II diabetes). Three different sets of variables we considered for hyperlipidemia: binary diagnosis of hyperlipidemia ("Vascular"), LDL level and statin use ("Vascular2"), and binary diagnosis of hyperlipidemia and statin use ("Vascular3"). HIV clinical factors included timing of ART initiation (from estimated date of HIV infection), duration of viral suppression, nadir CD4+ T count, pre-ART viral load, and ART regimen (given known effects of protease inhibitors on lipids and abacavir on NLRP3 inflammasome). Hazard ratios (HRs) are shown as risk per two-fold increase in individual cytokine levels at p<0.05 (*), as well as at a false discovery rate (FDR)-adjusted q < 0.05 (**). Bars represent 95% confidence intervals for each hazard ratio estimate using robust standard error.

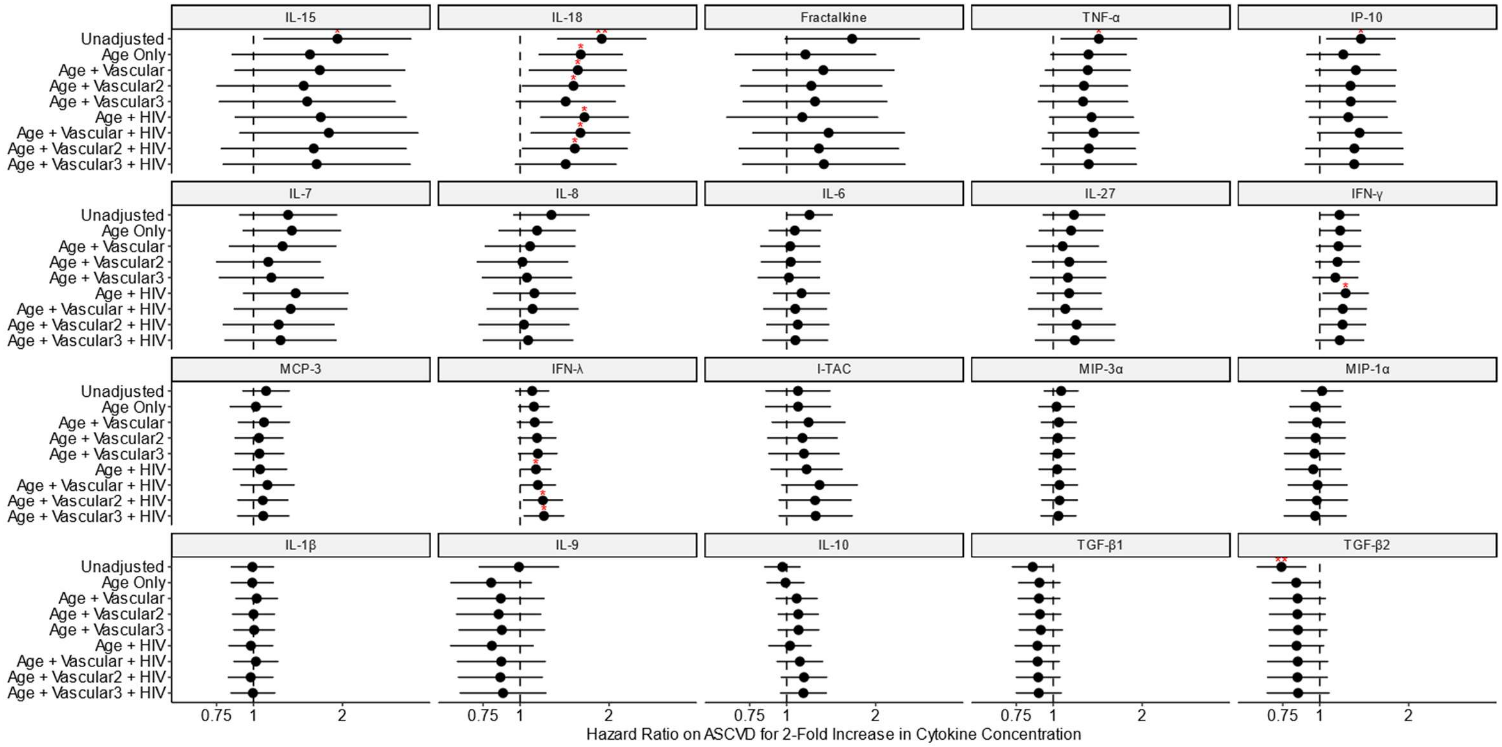

**eFigure 8. Unadjusted and adjusted hazard ratios of incident vascular event (VE), which include ASCVD and VTE diagnoses.** Linear unadjusted (black) and adjusted (grey) Cox proportional hazards models were used to assess the relationship between plasma cytokine concentrations (expressed as two-fold changes) and vascular outcomes. The adjusted models controlled for traditional vascular risk factors, including age (at plasma sample), sex, race/ethnicity, family history of vascular disease, tobacco use, and comorbidities (hyperlipidemia, hypertension, and type II diabetes). Cytokines that reached significance at the nominal p-value threshold ($p < 0.05$) or after false discovery rate (FDR) adjustment ($q < 0.05$) are marked with * and **, respectively. FDR correction was applied individually to each model. Bars represent 95% confidence intervals for each hazard ratio estimate using robust standard error.

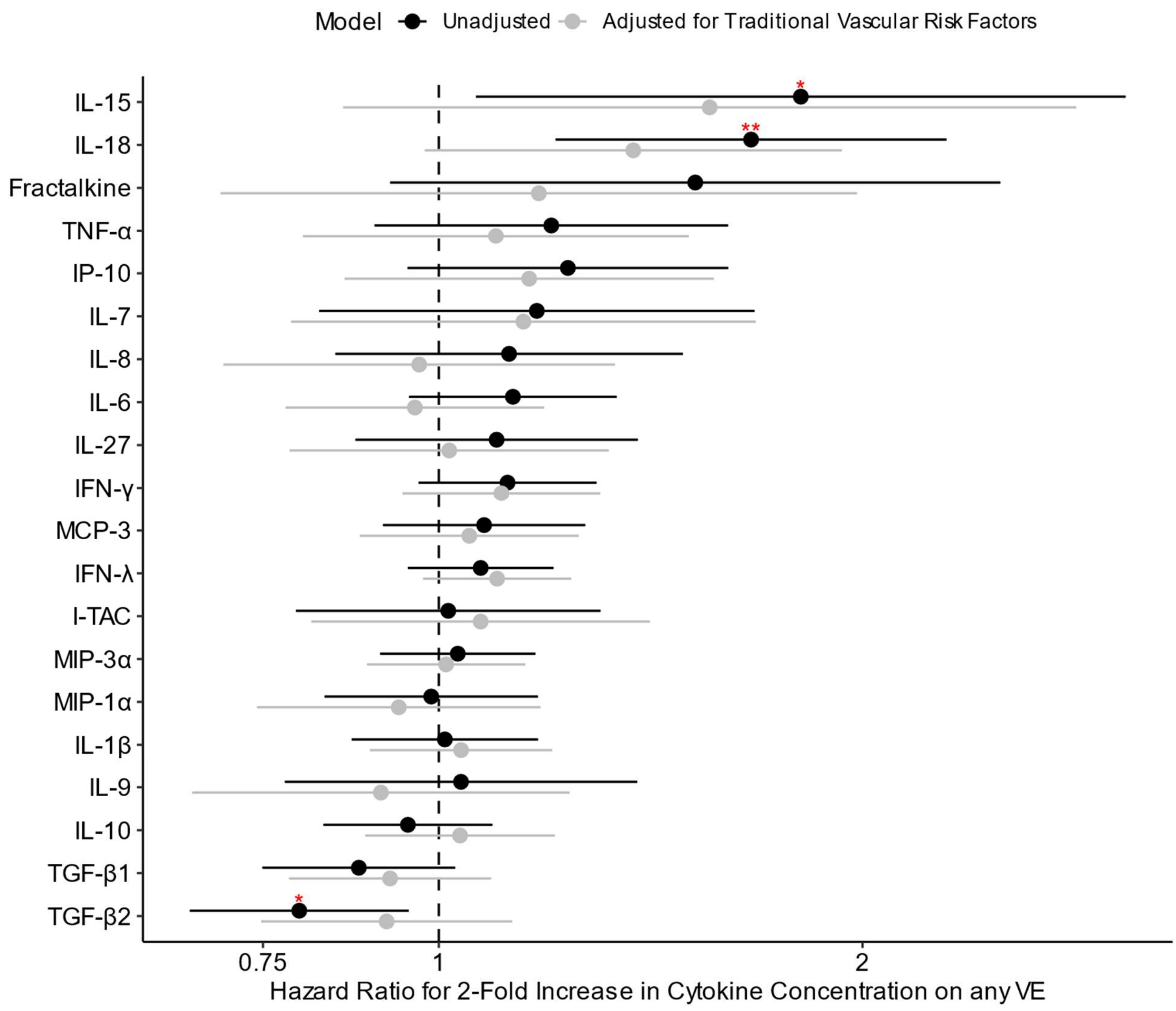

**eFigure 9. Unadjusted and adjusted hazard ratios for increase in plasma cytokine cluster medians, as predictors of ASCVD risk.** Linear Cox proportional hazard models were fit separately for each cytokine cluster, with unadjusted (black) and adjusted (grey) models presented. Hazard ratios (HRs) are reported per one unit increase in the median value of the cytokine cluster after log-transforming and scaling concentrations of each cytokine in the cluster to mean zero and unit variance. HRs are displayed for ASCVD risk across the eight clusters identified through unsupervised clustering (Figure 1). The adjusted models accounted for traditional vascular risk factors, including age at plasma sample collection, sex, race/ethnicity, hyperlipidemia, hypertension, type II diabetes, smoking status, and family history of vascular disease. Clusters that were significant at a nominal p-value level (p <0.05) or after false discovery rate (FDR) correction (q <0.05) are marked with * and **, respectively. FDR correction was applied individually to each model. Bars represent 95% confidence intervals for each hazard ratio estimate using robust standard error.

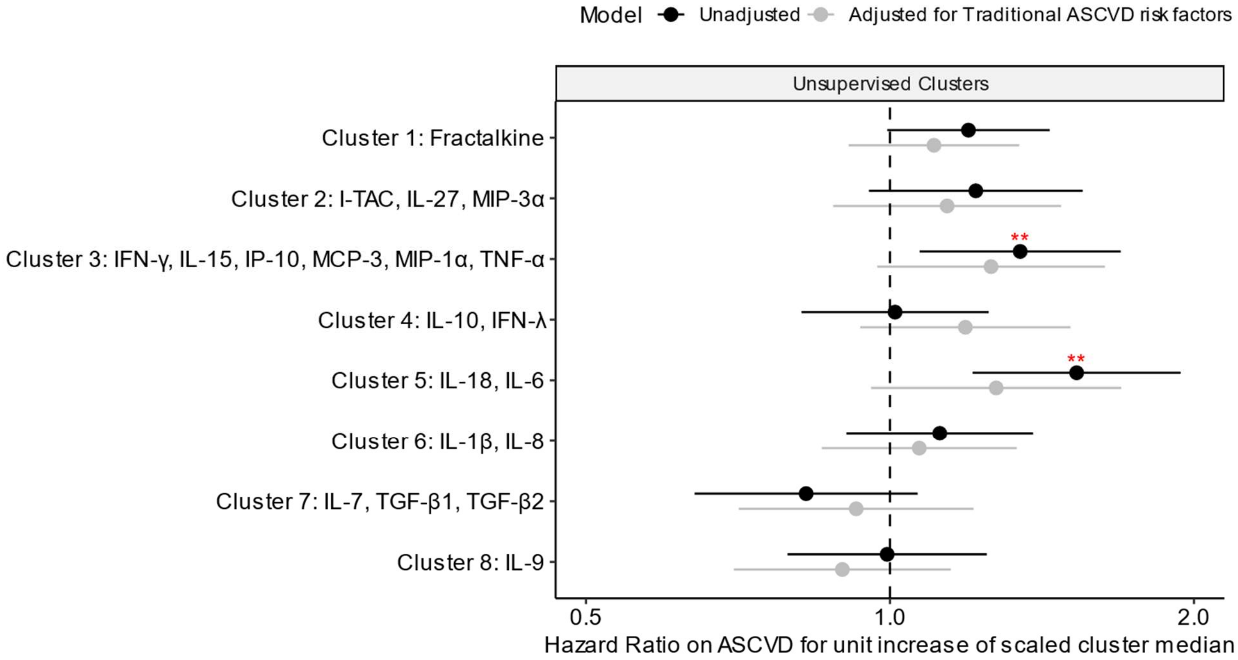

**eFigure 10. Higher levels of IL-18 but not TGF-β2 increase 5-year ASCVD risk in causal inference models.** The risk increases directly caused by doubling IL-18 (or TGF-β2) concentrations on 5-year ASCVD risk was estimated using a targeted maximum likelihood estimator. Both increase in relative risk (**A**) and absolute risk (**B**) are reported in the forest plots with respect to the baseline 5-year ASCVD risk for this cohort (approximately 3.86%). No association is indicated with a vertical dashed line. Bars represent 95% confidence intervals for each estimate. Models are adjusted for traditional ASCVD risk factors (age, sex, race/ethnicity, family history of vascular disease, tobacco use, comorbidities (diagnosis of hyperlipidemia, hypertension, or type II diabetes), and HIV clinical factors (timing of ART initiation, duration of viral suppression, nadir CD4+ T count, pre-ART viral load, and ART regimen). IL-18 showed a non-significant trend toward increased ASCVD risk, whereas TGF-β2 showed minimal association.

**A. (Relative Risk)**

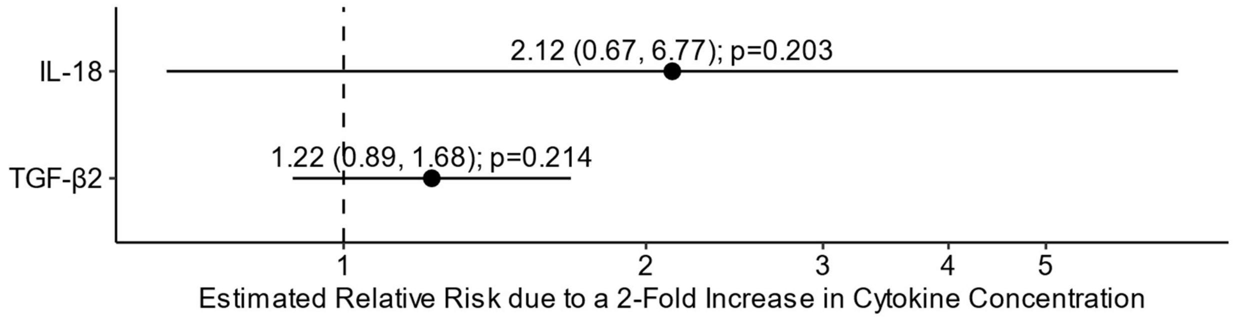


**B. (Absolute Risk)**

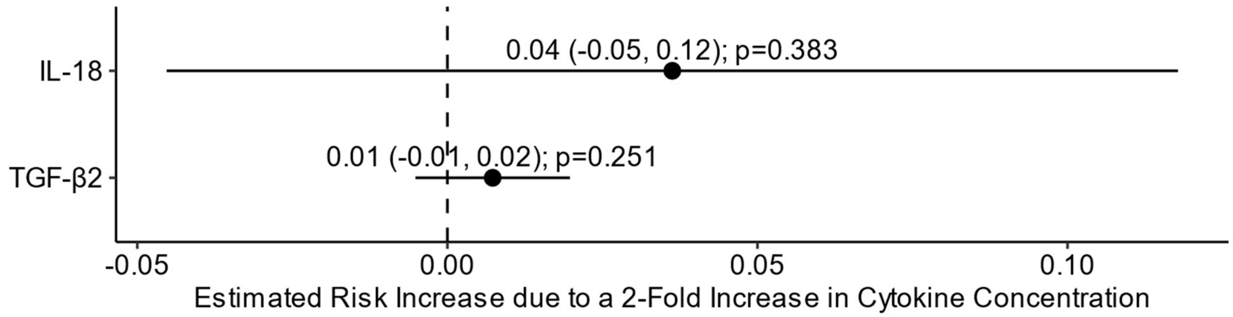

**eFigure 11. IL-18 as a persistent predictor of ASCVD risk after controlling for IL-6 and IL-1β.** IL-18 remained a statistically significant predictor of ASCVD risk, even after adjusting for IL-6 and IL-1β in the same model. Individual cytokine models are shown in the left panel (unadjusted) and right panel (adjusted). In these models, we compared the effect of IL-18 when adjusted for IL-1β and IL-6, both individually and together. For example, IL-18 remained significant after adjusting for IL-1β and IL-6 (p=3.5e-4 and p=0.001, before and after adjustment, respectively) in models not adjusted for traditional cardiovascular disease (CVD) risk factors. Adjusted models included covariates for traditional vascular risk factors – age at plasma sample, sex, race/ethnicity, hyperlipidemia, hypertension, type II diabetes, smoking status, and family history of vascular disease. Bars represent 95% confidence intervals for each hazard ratio estimate.

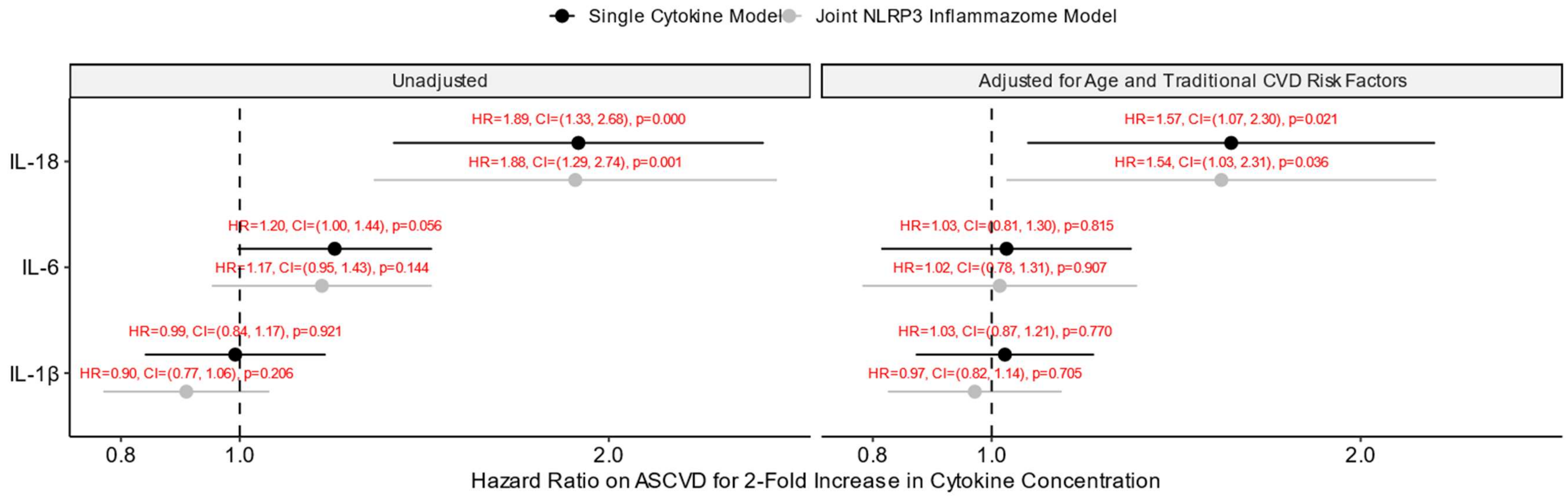
